# Non-pharmaceutical interventions and inoculation rate shape SARS-COV-2 vaccination campaign success

**DOI:** 10.1101/2021.02.22.21252240

**Authors:** Marta Galanti, Sen Pei, Teresa K. Yamana, Frederick J. Angulo, Apostolos Charos, Farid Khan, Kimberly M. Shea, David L. Swerdlow, Jeffrey Shaman

**Affiliations:** Department of Environmental Health Sciences, Mailman School of Public Health, Columbia University, 722 West 168th Street, New York, NY 10032; Medical Development and Scientific/Clinical Affairs, Pfizer Vaccines, 500 Arcola Road, Collegeville PA 19426; Patient and Health Impact, Pfizer Vaccines, 500 Arcola Road, Collegeville PA 19426

## Abstract

Nearly one year into the COVID-19 pandemic, the first SARS-COV-2 vaccines received emergency use authorization and vaccination campaigns began. A number of factors can reduce the averted burden of cases and deaths due to vaccination. Here, we use a dynamic model, parametrized with Bayesian inference methods, to assess the effects of non-pharmaceutical interventions, and vaccine administration and uptake rates on infections and deaths averted in the United States. We estimate that high compliance with non-pharmaceutical interventions could avert more than 60% of infections and 70% of deaths during the period of vaccine administration, and that increasing the vaccination rate from 5 to 11 million people per week could increase the averted burden by more than one third. These findings underscore the importance of maintaining non-pharmaceutical interventions and increasing vaccine administration rates.

## Introduction

The novel coronavirus SARS-COV-2, the causative agent of coronavirus disease 2019 (COVID-19), emerged in China during late 2019 and rapidly spread throughout the world. In March 2020, the World Health Organization (WHO) declared the COVID-19 a pandemic, and by January 2021 SARS-COV-2 had caused more than 100 million confirmed COVID-19 cases and 2 million deaths worldwide (*1*). A global effort to develop vaccines against SARS-COV-2 began early in 2020, but for most of that year the only options for slowing transmission were non-pharmaceutical interventions (NPIs), including stay-at home orders, encouraging the use of face masks, limiting in-person work and school, and social distancing.

In December 2020, the U.S. FDA granted emergency use authorization (EUA) for two COVID-19 vaccines that demonstrated safety and high efficacy in phase 3 trials: Pfizer/BioNTech BNT162b2 and Moderna mRNA-1273 (*2*). In light of limited supply, the Centers for Disease Control and prevention (CDC) recommended prioritizing vaccination per the following phases: 1a) healthcare workers and long-term care facility residents, 1b) priority essential workers and persons ≥ 75, and 1c) other essential workers, persons 65-75 and adults with pre-existing conditions (*3*). In the US, administration of BNT162b2 began on December 14, 2020 and administration of mRNA-1273 began on December 21, 2020. BNT162b2 and mRNA-1273 use mRNA technologies and require two doses administered 3 and 4 weeks apart, respectively, to reach full ∼95% efficacy (*4, 5*). By the end of 2020, the US has secured commitments for 400 million doses of these vaccines, which could be available for the US population by July 2021 (*6, 7*). However, by December 31,2020, fewer than 3 million doses had been administered, corresponding to 22.5% of the distributed doses at that time (*8*) and less than 15% of the anticipated target (*9*). During January 2021, the rate of vaccine administration increased. Presently, BNT162b2 is authorized for adults ≥ 16 years of age and mRNA-1273 for adults ≥ 18 years, but additional trials are being conducted to assess safety, immunogenicity and efficacy in children and pregnant women (*10, 11*).

### Model framework and analysis

In this analysis, we simulated and assessed the benefits of SARS-COV-2 vaccination in the US under varying levels of NPIs and differing vaccine administration and acceptance rates. Projections were made with a SEIRV (Susceptible-Exposed-Infected-Recovered-Vaccinated) compartmental model run in isolation for all 50 states and the District of Columbia (DC), in which the population was stratified by age and priority group. Specifically, we stratified each state population by years of age (0 - 4, 5 - 17, 18 – 49, 50 - 64 and ≥65), adult exposure status (essential workers (EW), healthcare workers (HC), other adults) and health risk status (presence or absence of one or more health risk factors for severe disease (RF)) (Figure 1A, Supplementary Tables S1 and S2). The model was parametrized using posterior distributions estimated with a separate, non-stratified metapopulation model iterated through January 10, 2021 (*12*) and later adjusted for age and population types (see Methods and Supplementary Table 2). Initial conditions and statistics for key epidemiological parameters are reported in Figure 1. The median estimated proportion of the state population susceptible (i.e. the population percentage not previously infected with SARS-COV-2) on January 10 was 65%, and varied across states as shown in Figure 1B. Initial susceptibility for the vaccine scenario projections were varied based on seroprevalence differences in the population (Figure 1C and Supplementary Table 2). The median national estimate of the time-varying reproduction number R_t_ was 1.78 on January 10; however, state-to-state heterogeneity of NPIs at that time is reflected in a broad distribution of R_t_ values ranging from 0.8 to 2.2 (Figure 1D).

**Figure 1:**
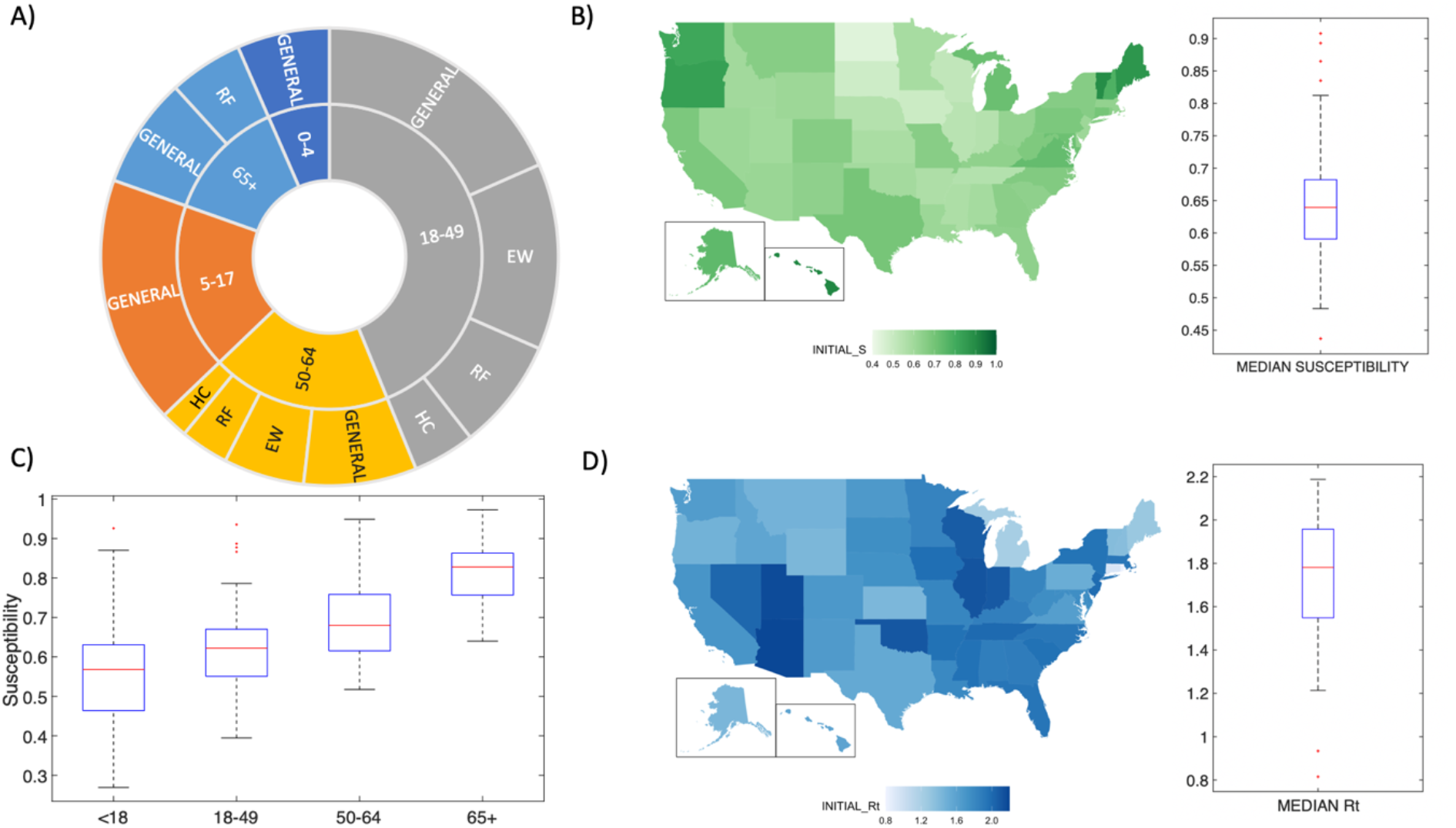
Initial conditions imposed on January 10, 2021. Panel 1A) represents the structure of the population in 12 groups classified by combination of years of age (0-4, 5-17, 18-49, 50-64, ≥65), exposure status (HC, EW, general population), and health risk factor (RF, non-RF). See Supplementary Table S1 and S2 for classification and overlapping factors. Panel 1B) shows the initial susceptibility as a fraction of each state population. The boxplot shows the median, interquartile range, and the full range of the distribution (outliers plotted in red) of the median values of population susceptibility for the 50 states and DC. Panel 1C) shows the distribution of susceptibility for different age groups among states. Children 0-4 years and 5-17 years are combined as available from CDC seroprevalence data (see Supplementary Table S2 for details). Panel 1D) presents a boxplot showing the median, interquartile range, and full distribution range (outliers plotted in red) for the median values of the time-varying reproduction number R_t_ on January 10, 2021 for the 50 states and DC.

All vaccination scenarios assumed 400 million doses (*6, 7*) distributed to the US population according to ACIP prioritization guidelines (*3*). We considered phases 1a, 1b and 1c completed 10 days after (first) vaccination of a target coverage number of individuals. Once the prioritization groups were vaccinated to target levels, vaccination was administered to other adults and children. The start date of the vaccination campaign was December 14, 2020, and, based on vaccination records (*8*), 5 million doses were administrated in the US through the first 3 weeks of the vaccination campaign. Doses were allocated to the 50 states and DC in proportion to state population size, and two doses of vaccine were administered to all vaccinated individuals 3.5 weeks apart (see Methods and Supplementary Tables S3 and S4 for details on vaccine modeling). Vaccination was administered regardless of prior history of infection and acted to prevent transmission to susceptible individuals. The impact of different scenarios was quantified in averted infections and deaths during the 15 months following January 10. The (mean) averted burden of infection (both ascertained and unascertained) was measured for each intervention scenario N_i_ with respect to the (mean) attack rate (AR) in reference scenario N_0_ (without vaccine and non-pharmaceutical interventions, see Supplementary Table S5) as:

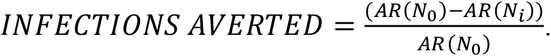

The same formula was used to quantify the averted deaths.

### Effect of NPIs under fixed vaccination scenarios

We first tested the effect of imposing different NPIs during the vaccination campaign. In this analysis, 5 million people received the first vaccine dose each week beginning week 4 of the campaign (Supplementary Table S4). We fixed the target coverage among different subpopulations at 80% for HC, 70% for risk groups (adults ≥65 and adults with RF), and 60% for other adults and children (up to available doses). The estimate of the time-varying reproduction number on January 10 (median R_t_=1.78) reflects reductions in opportunities for transmission due to NPIs, i.e., R_t_ is a reduction of the basic reproduction number (R_0_). Relaxing (strengthening) the NPIs would inflate (decrease) R_t_ and, in turn, the theoretical threshold for herd immunity. Estimates of the basic reproduction number for SARS-COV-2 in the United States vary across studies from 1.34 to 4 (*13,14*). Here we present the results for R_0_= 2.8; but also analyzed results for R_0_= 2.4 and R_0_= 3.2. Figure 2 compares the cumulative and averted burden of infection and death among 6 different NPIs scenarios characterized by different duration and strength of the NPIs imposed throughout the vaccination campaign: N0 is the limit scenario without intervention (NPIs or vaccination); N1 has vaccination but NPIs are completely relaxed on January 11, 2021; N2 maintains NPIs at initial levels then completely relaxes them upon completion of phase 1a; N3 relaxes NPIs in 3 steps upon completion of phases 1a, 1b, 1c; N4 first strengthens NPIs then relaxes in 3 steps after completion of phases 1a, 1b, 1c; and N5 maintains initial NPIs until (10 days after) 140 million people have initiated vaccination, then relaxes in 3 1-month steps. On average the 3 phases of vaccine prioritization in (*3*) were completed, respectively, 23, 66, and 154 days after January 11 (timing differed in each state due to population structure) and 140 million vaccinated were reached 193 days after January 11 (Figure 2A). Other NPI scenarios, including scenarios in which relaxation was triggered by time and not phase completion are described in Supplementary Table S5 and results are shown in Supplementary Figure S1.

**Figure 2:**
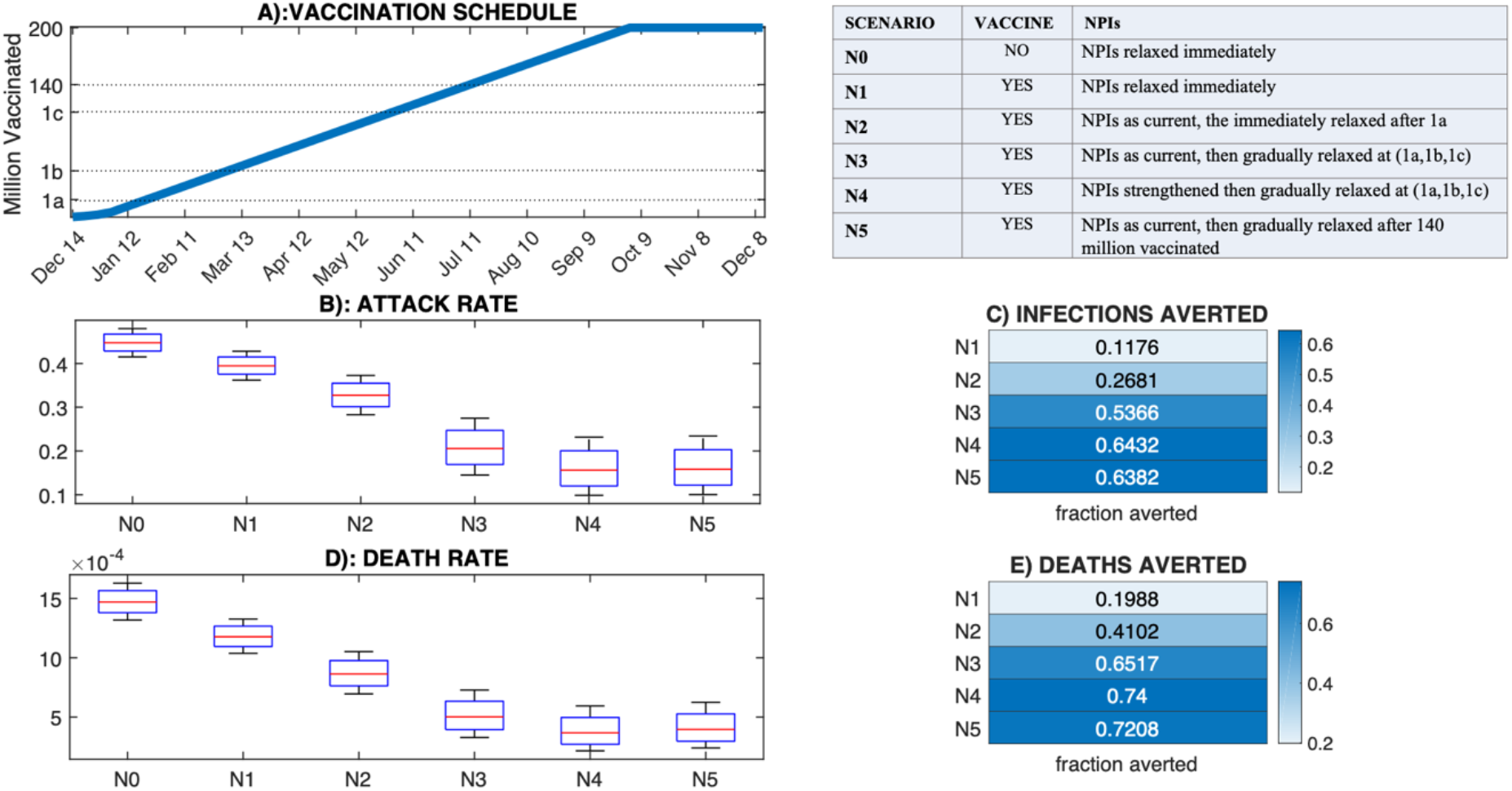
Effect of NPIs on infections and deaths with a fixed vaccination schedule. Panel 2A) shows the vaccination schedule (first doses) (see also Supplementary Table S4). Phase 1a), 1b), 1c) and 140 million vaccinated milestones are highlighted on the y-axis (the respective times on the x-axis do not include the additional 10 days required in the model for phase completion). The table panel summarizes the 6 NPI scenarios. Note that NPIs are eventually completely relaxed in all scenarios. Panels 2B) and 2C) show the attack rate and fractional reduction of infections for each scenario. Panel 2D) and 2E) show the death rate and fractional reduction of deaths for each scenario. Note, the attack and death rate do not include infections and deaths prior to January 11, 2021.

In Scenario N0 the cumulative attack rate in the overall population (for the period beginning January 11, 2021) was 44.8% and the cumulative death rate was 0.0015%. Adding vaccination (Scenario N1) yielded an attack rate of 39.5% and death rate of 0.0012%, an 11.8% reduction of infections and a 19% reduction of deaths relative to Scenario N0. Maintaining NPIs during the vaccination campaign allowed for much greater reductions: 20% to 60% of infections and 30% to 70% of deaths were averted depending on the strength of the NPIs maintained during the vaccination campaign. When NPIs were strengthened and gradually relaxed (N4) or maintained at initial levels for 6 months (Scenarios N5 and N6, N9 in Supplementary Figure S1) the attack rate in the population fell to roughly 15%. The more limited impact of vaccination in the absence of NPIs is due to the faster spread of SARS-COV-2: by the time phase 1a and 1b are completed in N1, 46% and 72% of the population is already immune (or deceased) by natural infection, whereas in N4 at the same time only 39% and 46% of the population have been infected (Figure 3). Without NPIs, vaccination had a weaker impact because 1) herd immunity was approached earlier during the campaign because the susceptible pool was diminished due to a high attack and 2) the rate of *effective vaccination* (vaccination of susceptible individuals) was slower due to a lower susceptible fraction. Results were robust across a larger set of scenarios and for different estimates of R_0_ (Supplementary Text S2 and Supplementary Figure S1), and were consistent at the state level. Among the 6 NPIs scenarios described here, N4 and N5 had the lowest attack rate in all states (Supplementary Figure S2). We also tested the sensitivity of the results to initial conditions (such as initial susceptibility) and vaccination setting (number of doses used, vaccine efficacy, and the consequence of vaccination protecting against disease instead of infection) (Supplementary Text S3, Supplementary Figures S3-S8). Though estimates of infections and deaths depended strongly on some of these varied parameters, the general finding indicating the strong effect of NPIs held.

**Figure 3:**
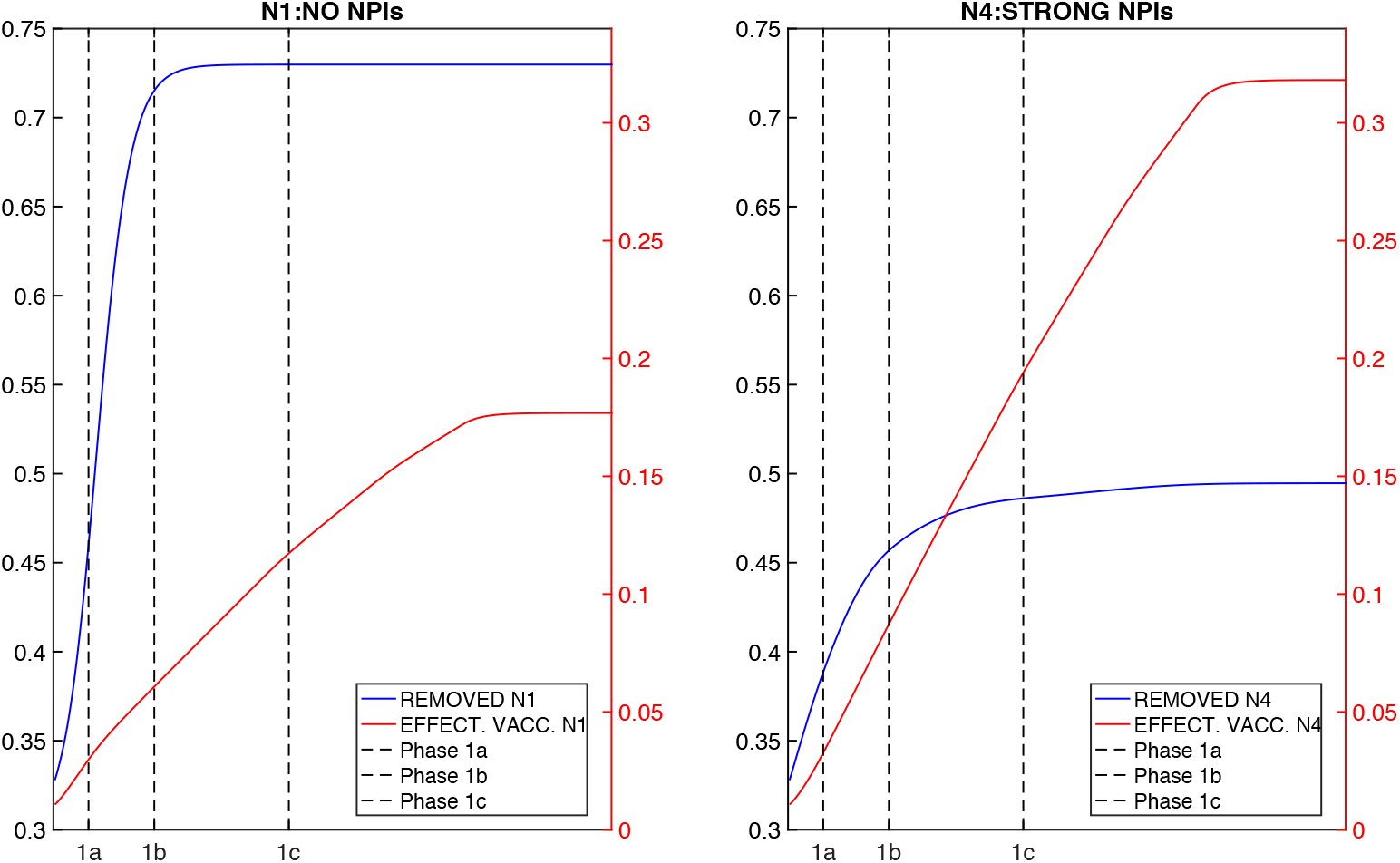
Effect of NPIs and vaccination on population immunity. Blue lines show the cumulative number of individuals no longer either susceptible or infected (i.e. recovered + deceased); red lines show the total effectively vaccinated (susceptible individuals who received the vaccine). The left panel shows the results from Scenario N1; the right panel shows the results from Scenario N4. Black vertical dashed lines mark the end of prioritization phases.

### Effect of vaccine deployment rate

A number of factors could affect the rate of vaccine administration in the coming months: availability of doses, distribution of doses, and management of the distributed stock by jurisdictions. We analyzed how variations in the rate of vaccine administration impact the cumulative infections and deaths averted due to vaccination. Specifically, we tested 6 vaccination schedules with 3, 5, 7, 9, 11 and 13 million people initiating vaccinations every week nationally. The 6 vaccination schedules were combined with 4 NPIs policies: a “NO NPIs” scenario with measures relaxed on January 11, 2021; a low distancing scenario (“LOW”) with NPIs completely relaxed after one month; an intermediate distancing scenario (“MED”) with NPI relaxation initiated after 1 month and gradually completed across 5 months; and a strong distancing scenario (“HIGH”) with measures first strengthened then gradually relaxed over 6 months. These scenarios correspond to scenarios N1, N7, N8, and N9 in Supplementary Table S5; all are characterized by a time-triggered relaxation of NPIs rather than a target-triggered relaxation in order to better isolate the effects of vaccination rate as phases 1a, 1b and 1c were reached at very different times across the 6 vaccination rates (e.g. the 3 prioritization phases were completed after 254 days for 3 million vaccinated/week and after 62 days for 13 million/week). The target coverage in each group remained the same as in the previous analysis: 80% for HC, 70% for risk groups, 60% for other adults and children up to availability.

Expanding the number of doses administered per week from 5 to 7 million averted 3 to 6% more infections and 4 to 8% more deaths with respect to Scenario N0; expanding from 5 to 11 million per week averted 9 to 16% more infections and 7 to 18% more deaths with respect to Scenario N0 (Figure 4). The effect of a faster deployment was stronger within scenarios characterized by weaker NPIs, because of the more rapid accumulation of infections in the first months. Results were robust to other estimates of R_0_ (Supplementary Text S4 and Supplementary Figure S9).

**Figure 4:**
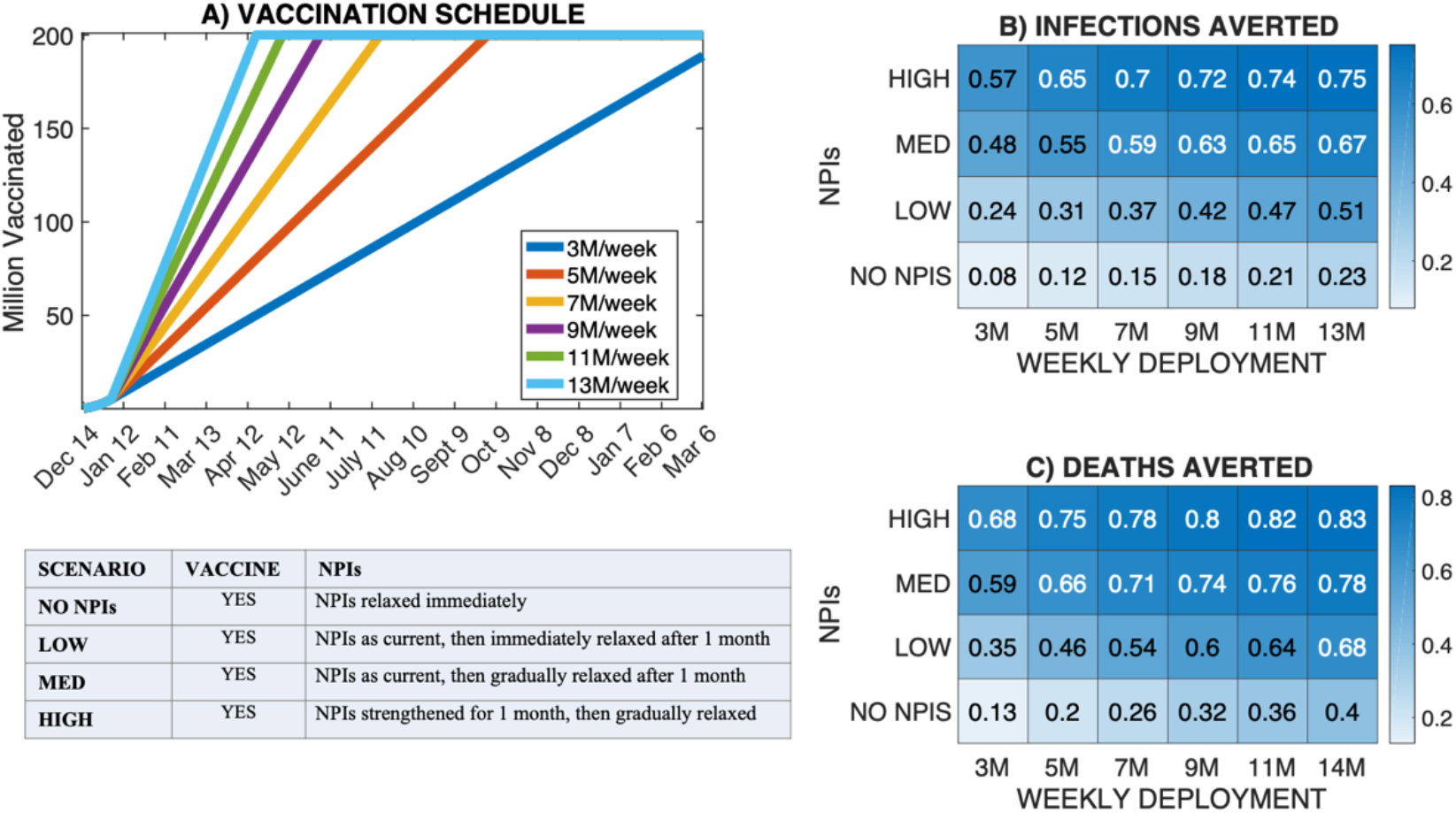
Effect of vaccine administration rate. Panel 3A) shows the vaccine administration timeline for the 6 vaccine deployment rates simulated. Panels 3B) and 3C) show the fractional averted burden of infections and deaths for each combination of administration rate and NPI scenario relative to Scenario N0. NPI scenarios NO NPIs, LOW, MED and HIGH correspond to Scenarios N1, N7, N8 and N9 of Supplementary Table S5.

### Effect of vaccine uptake

The third analysis examined the effects of vaccine uptake, specifically the percentage of each subpopulation able or willing to receive the vaccine (due to vaccine acceptance rates and difficulty accessing vaccination facilities), on population outcomes. Here, we assumed 5 million doses distributed per week beginning January 11, 2021. We assessed the effect of uptake by comparing the cumulative infections and deaths for the same 4 NPIs scenarios, N1 (NO NPIs), N7 (LOW), N8 (MED), N9 (HIGH), considered in the previous analysis. Baseline coverage, *c*, remained 80% among HC, 70% among individual at risk and 60% among other adults and children up to availability. We then tested different uptake levels by increasing or decreasing the coverage of all groups by the same percentage (Scenarios c_0.5_, c_0.75_, c_1.2_ are obtained multiplying the baseline uptake c, respectively, by 0.5, 0.75 and 1.2). Additionally, for Scenario c_99_ target coverage is set to 99% for the whole population, and for Scenario c_R_ target coverage is increased to 99% only for higher risk groups and kept at baseline for other groups.

Given 400 million doses, the maximum percentage of the overall population that could be vaccinated was 61.5% overall, therefore c, c_1.2,_ c_99_ and c_R_ reached the same cumulative coverage (see Figure 5). The effect of a uniform increase or decrease in uptake across all groups was moderate, whereas a stronger impact on deaths averted was seen when increasing uptake solely for higher risk groups, consistent with other recent findings (*15*). Specifically, uniformly doubling uptake from 32% to 64% of the population averted 2 to 4% more infections and 3% to 5% more deaths with respect to the no intervention Scenario N0 when some level of NPIs were also imposed. In the NO-NPI scenario (N1) doubling the uptake from c_0.5_ only averted 1% more infections and did not increase averted deaths. On the other hand, Scenario c_R_ averted 4% more deaths with respect to N0 than the baseline Scenario c with equal cumulative coverage (Figure 5).

**Figure 5:**
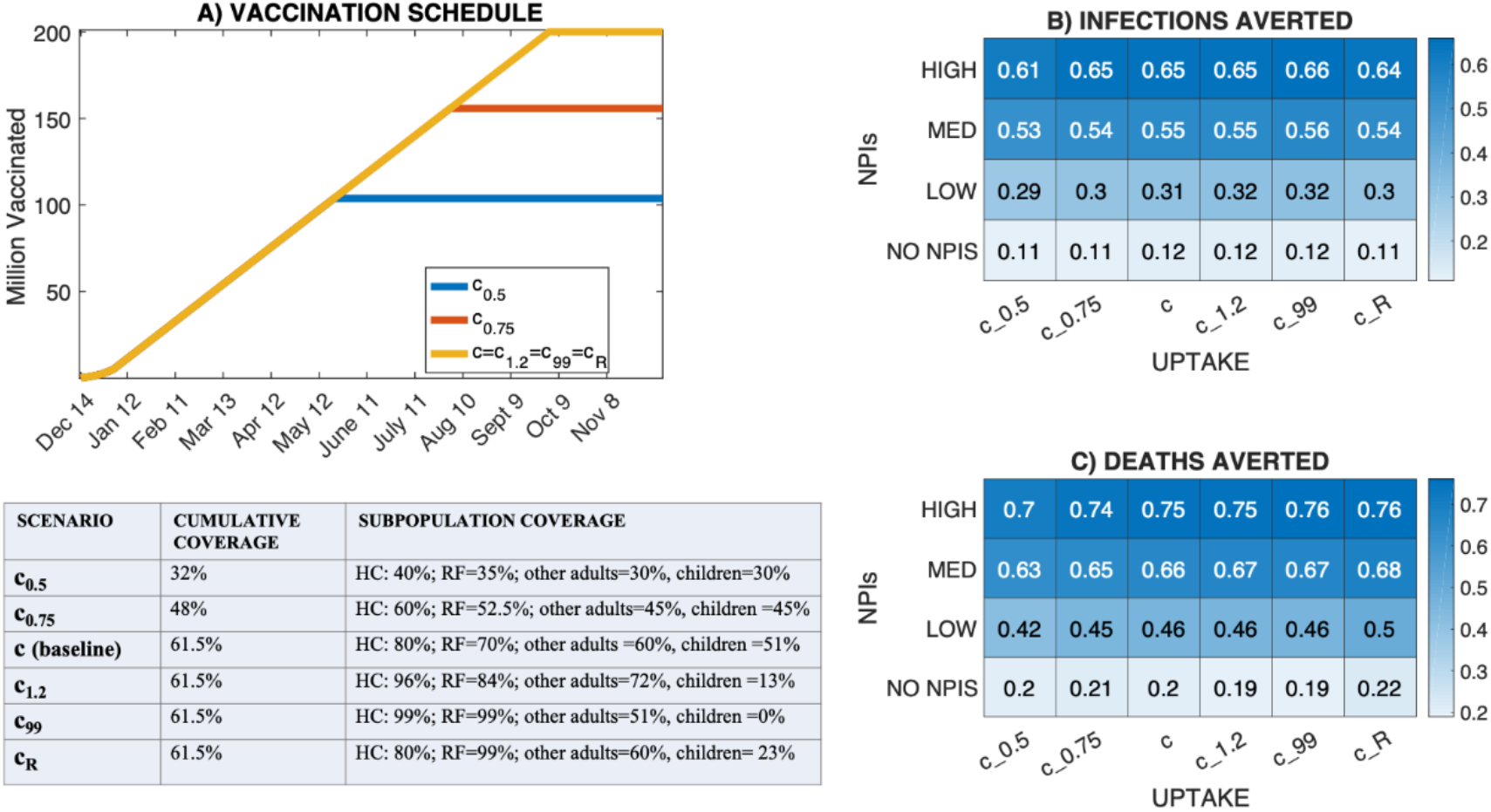
Effect of vaccine uptake on infections and deaths. Panel 5A shows the vaccine distribution timeline for the different vaccination uptake scenarios (c_0.5_, c_0.75_, c, c_1.2_, c_99_, c_R_). Panels 5B and 5C show the fractional averted burden of infections and deaths for each specific combination of vaccination uptake and NPIs scenarios relative to baseline Scenario N0. NPIs scenarios NO NPIs, LOW, MED and HIGH correspond to scenarios N1, N7, N8 and N9 of Supplementary Table S5.

Two processes appear to explain this result: 1) vaccine impact is greater in the first months of administration when fewer natural infections have occurred, but, at that time, demand exceeds vaccine availability, and 2) increased coverage in not-at-risk groups has the effect of delaying the administration to lower priority but more at-risk-groups, which, with the exception of LTCF residents, were included in phases 1b and 1c. In Scenario c_0.5_ only 35% of the population at risk was vaccinated, but phase 1b and 1c started 2 weeks and 1 month, respectively, earlier than in Scenario c (Supplementary Figure S10). This earlier administration to the 1b and 1c groups partially offset the lower vaccinated proportion. The averted infections for c_99_ varied minimally even when increasing the total doses from 400 to 600 million, suggesting that increasing the overall coverage is not very effective without increasing the weekly vaccination rate (Supplementary Text S5). Results were robust to different estimates of R_0_ (Supplementary Figure S11).

## Discussion

The recent advent of safe and efficacious SARS-COV-2 vaccines could help end the pandemic. However, even in the most optimistic scenario, administering full vaccination with either BNT162b2 or mRNA-1273 to most of the population will take many months to complete, due to time required for production, distribution, and administration of two doses. According to the agreements stipulated in December 2020 between the US government and the vaccine manufacturers (*6, 7*), the US has purchased enough doses of BNT162b2 and mRNA-1273 for full vaccination of more than 60% of the population. An additional 100 million doses of both BNT162b2 and mRNA-1273 were recently contracted by the US government, other vaccine candidates are currently undergoing or completing phase 3 trials, and negotiation with manufacturers is ongoing (*16*); thus, it is possible that additional vaccines and additional doses of BNT162b2 and mRNA-1273 may contribute to increased vaccine coverage in the coming months.

Here we performed an analysis to test how the impact of the vaccination campaign (with a fixed total of 400 million doses) depend on 3 factors: (1) the NPIs imposed during vaccination, (2) the rate at which doses are administered, and (3) the vaccine uptake within subpopulations differing by age, exposure status and health risk status. The strongest modulator of the impact of vaccination, measured by averted infections and deaths for a broad range of realistic scenarios was the enforcement of NPIs throughout the vaccination campaign. With stronger NPIs, virus transmission slows, allowing vaccination of more susceptible people prior to infection. Overall, the vaccination campaign over the next several months has the potential to prevent infection of 20 to 40% of the US population; however relaxing NPIs before attaining adequate vaccine coverage could result in infection of those individuals and further hospitalizations and mortality. In the scenario in which all NPIs were immediately relaxed 4 weeks into the vaccination campaign, the averted infections were only 14%-27% of the number averted in the strongest NPIs scenarios, depending on the estimate of R_0_. When NPIs were maintained for a long time, hundreds of thousands of deaths were averted at the national level. These findings underscore the importance of maintaining NPIs during the vaccination campaign. However, we are now over one year into the pandemic; exhaustion and the economic toll of the pandemic cannot be discounted by policy makers in evaluating the extent and duration of the NPIs to be enforced during the next months.

The administration rate of the vaccine also had a strong impact in our analysis: increasing weekly vaccinations from 5 to 11 million, while keeping the cumulative availability fixed, reduced deaths by 17 to 20% with respect to the no intervention scenario across different estimates of R_0_. Increasing the speed of vaccine administration was particularly important for scenarios with reduced levels of NPIs. It is therefore essential to increase efforts to produce, distribute, and administer the vaccine.

In the first weeks of the US vaccination campaign, only 20% of distributed doses were administered. By January 28, 2021, that percentage increased to about 50% (*8*). Several factors need to be optimized: coordination between the federal government and individual states, management of the vaccine stocks by jurisdictions, operation of vaccination sites including coordination of personnel and strategies for facilitating population access, and protocols to assure that vaccine doses are not wasted.

In our analysis, the effect of vaccine acceptance on the overall averted disease burden of the vaccine campaign was limited. This was due to two factors: 1) with a fixed administration rate of 5 million doses per month, the vaccination campaign had a greater impact in the first months, when vaccine demand was greater than availability, even for low vaccination uptake scenarios; and 2) the prioritization order did not place risk groups with higher mortality first in line for vaccination. Therefore, in a low uptake scenario the non-risk groups were processed faster, allowing the risk groups to be vaccinated earlier, albeit with a lower uptake. This result, however, has to be evaluated within the limitations of our approach: first, we only considered mortality and infection, whereas the pandemic has an impact also on Quality-Adjusted Life Years (QALYs) and Disability-Adjusted Life Years (DALYs), as well as occupational hazard and social disparity. Second, even though we characterized healthcare workers as having more work contacts than other adults, we didn’t characterize those contacts as more likely to be with infected individuals. Therefore, the averted burden of infections with the current prioritization could be underestimated.

In our simulations, we evaluated full vaccination with BNT162b2 and mRNA-1273, which both require two doses, as recommended by the FDA (*17*). In light of recent consideration of reducing the number of doses from 2 to 1 until adequate vaccine is available, we additionally examined how a single dose campaign with BNT162b2 and mRNA-1273 could modify the impact of vaccination. The one-dose campaign, which doubled the weekly vaccination rate but with 90% vaccine efficacy, averted up to 12% more infections and 15% more deaths across different NPI scenarios relative to the no-intervention scenario. However, caution is needed when interpreting model results for single dose administration, as it is unknown whether immunity would last as long as with two doses. Indeed, other vaccines (e.g. Tetanus and Hepatitis B vaccines) require additional doses administered within a short time in order to boost and ensure a robust and durable adaptive immune response (*18*).

Our analysis has several limitations. The high dimensional model is sensitive to the choice of parameters and initial conditions. We tested finding sensitivity to several model parameters and even though the numbers of averted infections and deaths varied, sometimes largely, as with initial population susceptibility (see Supplementary Figure S3), the overall conclusions of our analyses held.

In our study we model vaccine as 95% efficacious (after 2 doses) in preventing infection and, as a consequence, transmission of SARS-COV-2. However, the primary end points of the phase 3 trials for BNT162b2 and mRNA-1273 were efficacy against confirmed COVID-19 disease in vaccine recipients, not infection. There is some evidence (*19*) indicating vaccination reduces asymptomatic infection rates. However, at the moment observations are limited and more data are needed to distinguish vaccine efficacy in preventing infection and disease. Should the vaccines prove efficacious in preventing disease but not infection, the impact of vaccination on overall attack rate would likely be more limited than the effects shown here (see Supplementary Text S3 and Supplementary Figure S8).

In this analysis, we also did not account for waning natural or vaccine-induced immunity or the emergence and dissemination of SARS-CoV-2 variants for which vaccines may be less efficacious (*20*) (however, we did test the sensitivity of results to lower efficacy of the vaccine (see Supplementary Text S3)). Evidence of re-infections with SARS-COV-2 has been reported around the world (*21, 22, 23*); however, more data are needed to understand the effect and time scale of these events at the population level. Should immunity prove to be short-lived, vaccination may need to be repeated every year or every few years for adequate coverage. A different model structure, accounting for loss of immunity, would be needed to quantify the burden of infection and deaths in this instance.

Overall, our findings indicate that vaccines can have a profound impact on the pandemic including prevention of many deaths. The public health objective is to vaccinate as many people as possible prior to infection. To do so, production, distribution and administration of vaccine must be accelerated and NPIs kept in place until enough doses are delivered to prevent sustained community transmission.

## Data Availability

Inference data and model output are available on the Github repository https://github.com/shaman-lab.

## Author Contributions

JS, MG, FA conceived the study, MG, FA, DS, AC, KS, FK, SP, TKY, JS designed the study. MG, SP, TKY performed the analysis, MG drafted the manuscript, all authors revised and reviewed the manuscript.

## Data and materials availability

Inference data and model output are available on the *Github* repository https://github.com/shaman-lab.

## SUPPLEMENTARY MATERIAL

### Supplementary Text S1: Methods

#### 1.1 Model Description

The principal model is a single location Susceptible-Exposed-Infected-Recovered-Vaccinated (SEIRV) compartmental structure run in isolation for each state in the US. We accounted for 5 age groups and 4 population types for the adult categories: healthcare workers (HC), essential workers (EW), individuals with pre-existing health conditions known to be risk factors for severe disease (RF) and persons other than HC, EW and RF (general) (see Table S1). We assume the 12 groups are mutually exclusive. The compartmental model is a modification of the structure that our group has used for inference and forecast for multiple infectious diseases, including influenza (*24*).

**Table S1:** Population stratification.

|  | AGE GROUPS | POPULATION-TYPE |
| --- | --- | --- |
| 1 | 0-4 | general |
| 2 | 5-17 | general |
| 3 | 18-49 | general |
| 4 | 18-49 | HC |
| 5 | 18-49 | EW |
| 6 | 18-49 | RF |
| 7 | 50-64 | general |
| 8 | 50-64 | HC |
| 9 | 50-64 | EW |
| 10 | 50-64 | RF |
| 11 | $\geq 65$ | general |
| 12 | $\geq 65$ | RF |

Broadly, the model distinguishes between reported infections *I* and unreported infections *U*, with the former being more contagious and likely transmit virus. Natural infection (both reported and unreported) is assumed to confer permanent immunity. Vaccines are administered according to a specific prioritization calendar, and individuals are vaccinated regardless of previous infection record. However, only susceptible individuals receiving the vaccine transition to the Vaccinated compartment. The vaccine is administered in two doses 3.5 weeks apart to all recipients. Vaccine efficacy is modeled as a 3-step function with values:

*ϵ*_1_ until 12 days after first dose,

*ϵ*_2_ 12 days after first dose to 1 week after second dose

*ϵ*_3_ after 1 week from second dose.

Estimates of vaccine efficacy and further details on vaccination are given in section 1.2.3 below and Supplementary Table S3.

The complete stratified model for each of the 50 states and DC, s=1:51, is:

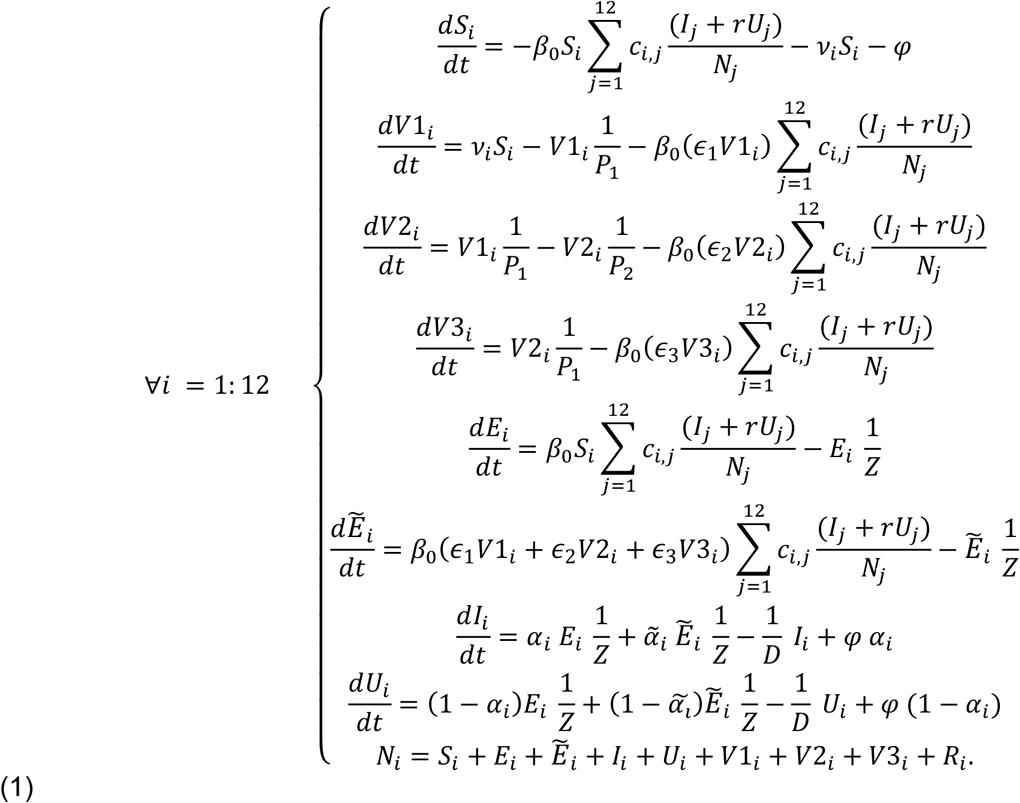

For each subpopulation *i, S*_*i*_, E_*i*_, 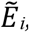, *I*_*i*_, *U*_*i*_, *V1*_*i*,_ *V2*_*i*,_ *V3*_*i*,_ and *R*_*i*_ represent the susceptible, exposed, exposed vaccinated, infected reported, infected unreported, vaccinated (in thse 3 phases) and recovered populations, *D* the duration of infectious period, Z the latency period, and *N* the population size. We distinguish between Exposed and Exposed Vaccinated to allow for a different probability of developing reported infection: *α*_*i*_ and 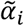 the ascertainment rate of infection, respectively, for unvaccinated and vaccinated individuals. Other parameters are: *φ* the travel-related importation of SARS-COV-2 into the model domain, *v*_*i*_ the vaccination rate, and *r* the decreased probability of transmission for unreported infectious individuals. We allow the transmission rate to vary through specific age-dependent contact rates *c*_*i, j*_ between individuals in age groups *N*_*i*_ and *N*_*j*_, such that the full transmission term is 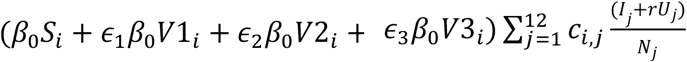. In the present model, we do not consider waning immunity and the possibility of reinfection. Deaths were calculated from model output using a Case Fatality Rate (CFR) specified by age and pre-existing condition based on case report CDC data (see Supplementary Table 2 for details).

One hundred ensemble projections were simulated in each state for 450 days starting from initial conditions estimated on January 10, 2021. Projections for each state were initialized using posterior estimates derived from a separate metapopulation model-Bayesian inference system, described in Section 1.2.1.

#### 1.2 Initialization of parameters

Our strategy for defining the distributions of parameters and variables in system (1) is based on the following steps:

1. We estimated the *population-level* distribution (interquartile range) of the epidemiological parameters on January 10, 2021 in each State with a non-stratified metapopulation model-Bayesian inference framework.
2. We combine the *population-level* estimates with information on population structure and age-specific infection and seroprevalence rates to stratify the initial conditions by age and population type (see Supplementary Table S2 for details on specific parameters and variables).
3. We initialized system (1) with the stratified distribution of parameters and initial conditions. For each state, we ran 100 simulations, each with initial conditions and parameters randomly drawn from the interquartile of the estimated distributions.

##### 1.2.1 Inference model

To perform the inference in step 1) we used a county-scale metapopulation model in which transmission of SARS-CoV-2 is simulated within and between each of the 3142 counties of the US (*12*). The subpopulations of each county are linked by documented rates of inter-county commuting and random travel. The metapopulation model has a SEIR structure featuring the same distinction between reported and unreported cases, but without the age/exposure stratifications listed in Table S1. The metapopulation model is combined with the ensemble adjustment Kalman filter (*25*), which assimilates case observations (reported infections) and iteratively estimates the time-evolving distribution of unobserved parameters and state variables. Details on the inference procedure can be found in (*12*). Using this framework, we obtained population level (i.e., unstratified) estimates of initial conditions on January 10, 2021 (e.g., the time-varying reproduction number *R*_*t*_, initial susceptibility, ascertainment rate, etc.).

##### 1.2.2 Mapping to stratified estimates

We combined the inferred parameters with information on population structure, age specific seroprevalence estimates and published age-specific infection reporting rates as described in Supplementary Table S2.

**Table S2:**
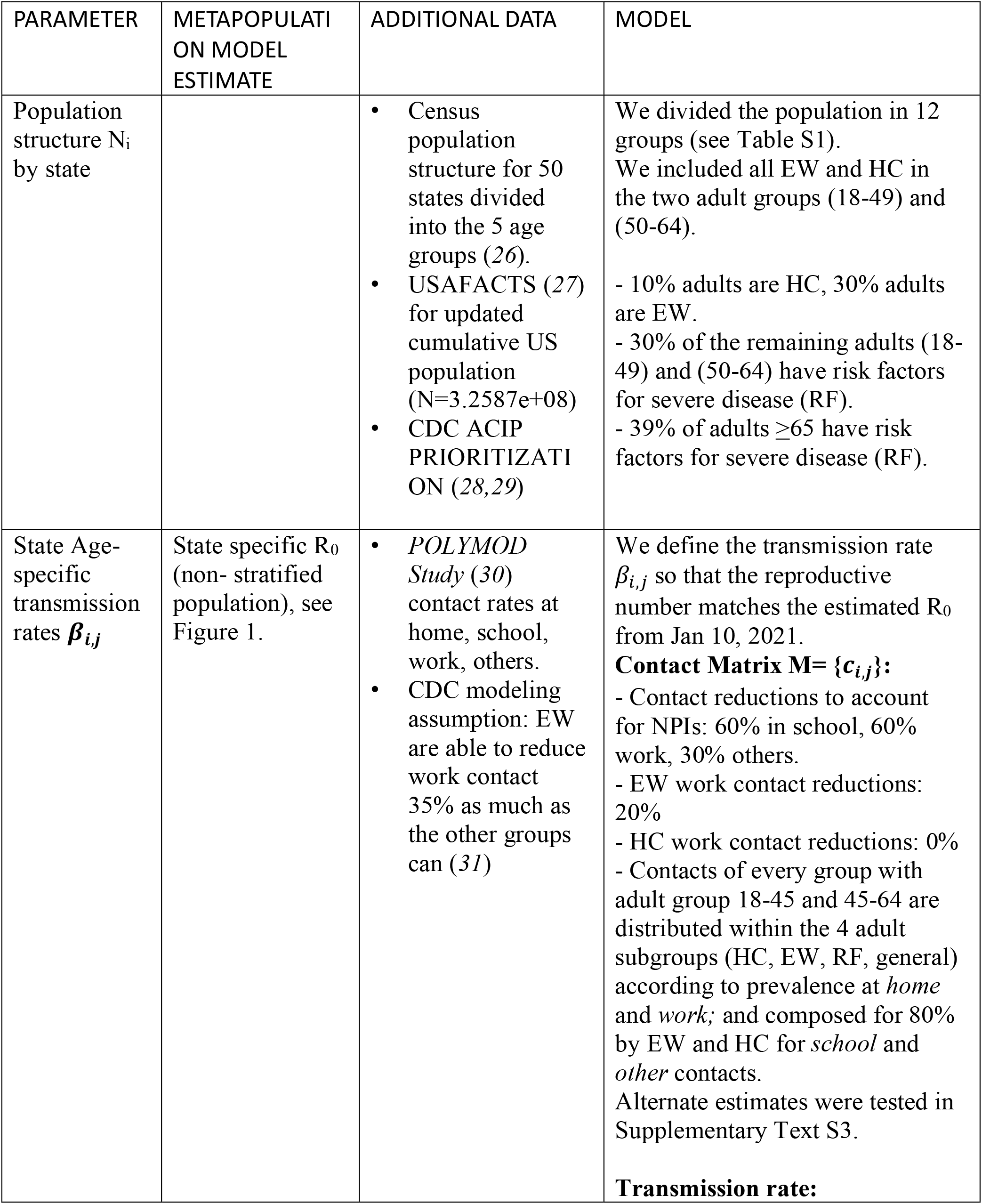
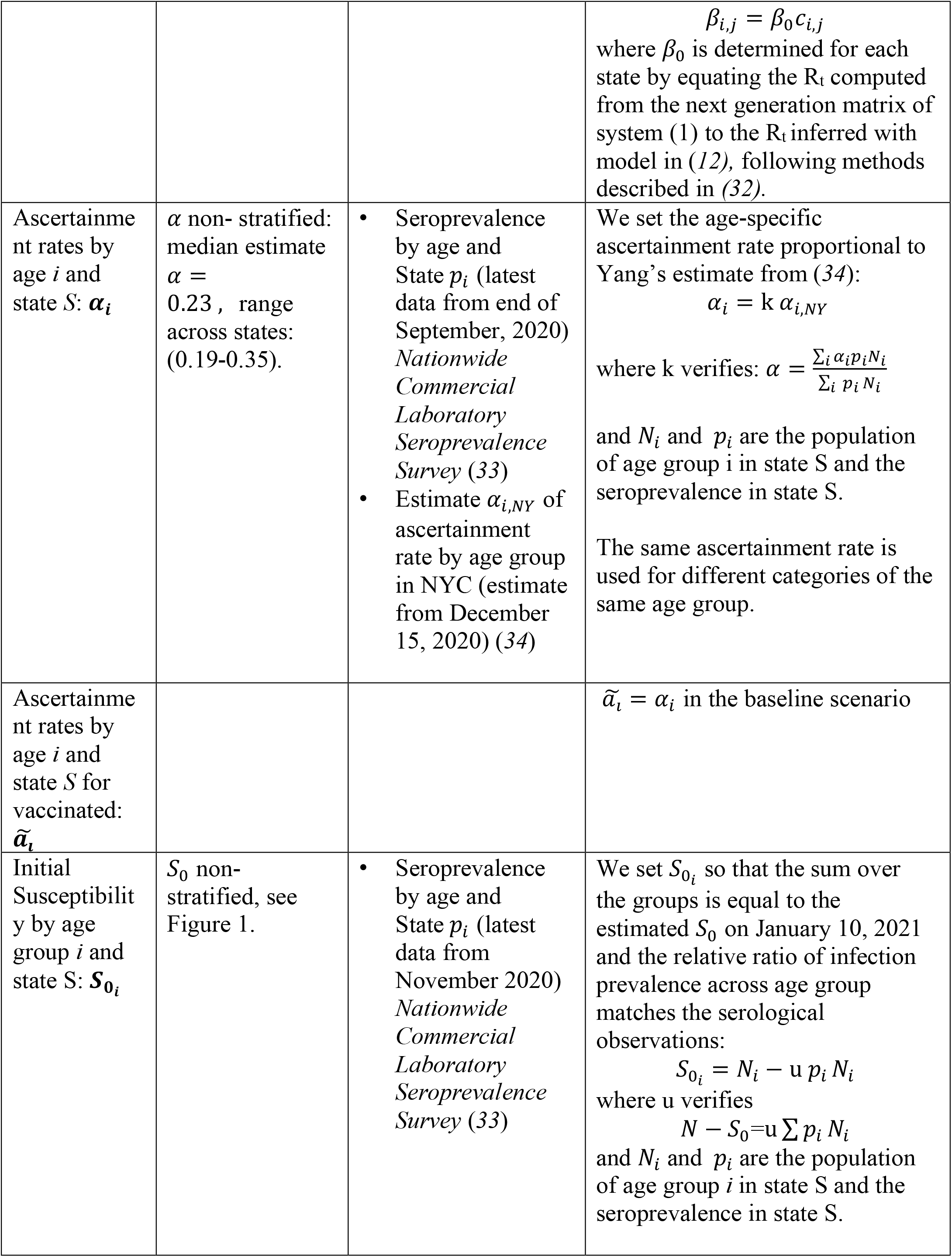
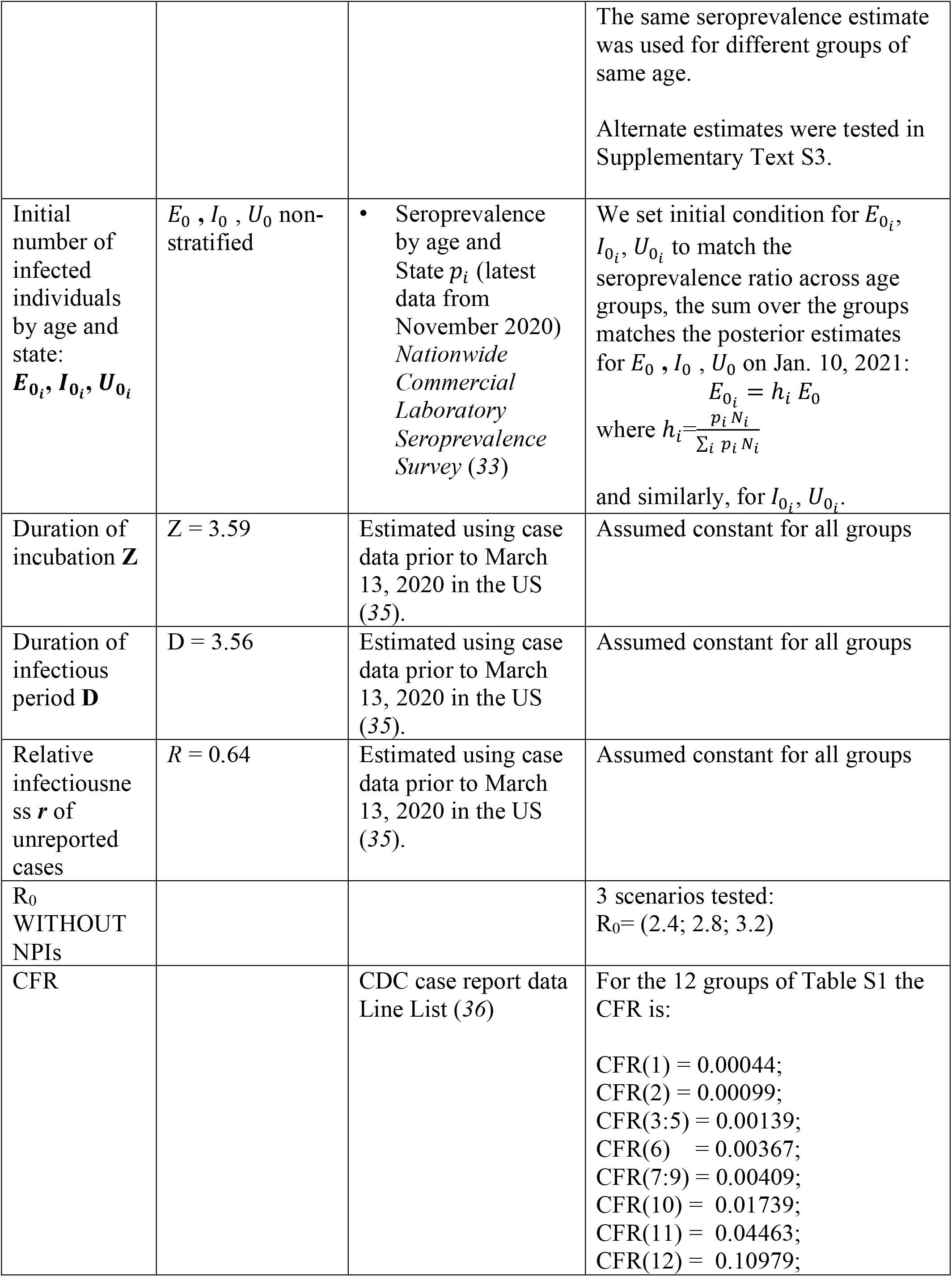
Age- and population-specific parameter specifications. All parameters and variables are state-specific; we omit the subscript _S=1:51_ for readability. *Estimated* parameters were inferred using the model-inference framework (*12*) (Section 1.2.1) and incidence data at the county level through January 10, 2021.

##### 1.2.3 Vaccination-related parameters

Vaccination administration in the US began on December 14, 2020. We assumed 5 million doses administered cumulatively in the first 3 weeks based on inoculation records (*8*). In subsequent weeks, beginning January 4, 2021, we assumed 5 million people vaccinated nationally for each subsequent week (except for the administration rate analysis in which weekly rates varied from 3 to 13 million vaccinated). We assumed both the BNT162b2 and mRNA-1273 vaccines required 2 doses and that complete efficacy was reached 1 week after the second dose. The time between first and second dose is 3 weeks for BNT162b2 and 4 weeks for mRNA-1273 (*4, 5*). Here, we averaged these two interval times and assumed that each individual received the second dose 3.5 weeks after the first dose. Weekly doses were assumed to be distributed uniformly over 7 days. Vaccine parameters and the administration timeline for the baseline scenario are detailed in Table S3. Vaccination in the simulations was administrated according to the following prioritization order based on (*3*):

1. Phase 1a: Healthcare workers (∼20 million) & long-term care facility (LTCF) residents (∼3 million).
2. Phase 1b: Front Line Essential Workers (∼30 million) & adults ≥65 with RF (∼14 million)
3. Phase 1c: Other Essential Workers (∼30 million), adults ≥65 (∼26 million) & adults with RF (37 million)
4. Other adults (∼86 million)
5. Children (∼79 million). Although both the BNT162b2 and mRNA-1273 vaccines are not currently recommended for children, in our analysis we hypothesize that by the end of the vaccination campaign vaccine will be recommended to all age groups.

In each of the 5 phases, individuals were immunized up to a target coverage (baseline target coverage was 80% for HC, 70% for adults ≥65 and adults with RF, and 60% for others.). Phases were considered completed for the purposes of NPI relaxation (used in some scenarios) 10 days after reaching target coverage (for first vaccination). The weekly timeline of baseline scenario vaccination is shown in Table S4.

**Table S3:** Vaccination parameters.

| PARAMETER | SOURCE | ESTIMATE |
| --- | --- | --- |
| VACCINE EFFICACY | Kaplan Meier curve for efficacy estimate (4) | $\epsilon_1$: 0% for 12 days after 1<sup>st</sup> dose<br>$\epsilon_2$: 90% from 12 days after 1<sup>st</sup> dose to 7 days after 2<sup>nd</sup> dose<br>$\epsilon_3$: 95% from 7 days after 2<sup>nd</sup> dose<br>Alternate estimate tested in Supplementary Text S3 |
| TIME BETWEEN DOSES | Pfizer-BNT162b2: 3 weeks (4)<br>Moderna-mRNA-1273: 4 weeks (5) | 3.5 weeks |
| TOTAL DOSES AVAILABLE | (6, 7) | 400 MILLION (enough to fully vaccinate 200 million people) |

**Table S4:** Weekly (first) vaccine distribution timeline (in millions). Weeks are identified by the first day of the week. Weekly doses are distributed uniformly over 7 days.

| WEEK | PEOPLE INITIATING VACCINATION | CUMULATIVE POPULATION INITIATING VACCINATION |
| --- | --- | --- |
| 14-Dec | 1 | 1 |
| 21-Dec | 1.5 | 2.5 |
| 28-Dec | 2.5 | 5 |
| 4-Jan | 5 | 10 |
| 11-Jan | 5 | 15 |
| 18-Jan | 5 | 20 |
| 25-Jan | 5 | 25 |
| 1-Feb | 5 | 30 |
| 8-Feb | 5 | 35 |
| 15-Feb | 5 | 40 |
| 22-Feb | 5 | 45 |
| 1-Mar | 5 | 50 |
| 8-Mar | 5 | 55 |
| 15-Mar | 5 | 60 |
| 22-Mar | 5 | 65 |
| 29-Mar | 5 | 70 |
| 5-Apr | 5 | 75 |
| 12-Apr | 5 | 80 |
| 19-Apr | 5 | 85 |
| 26-Apr | 5 | 90 |
| 3-May | 5 | 95 |
| 10-May | 5 | 100 |
| 17-May | 5 | 105 |
| 24-May | 5 | 110 |
| 31-May | 5 | 115 |
| 7-Jun | 5 | 120 |
| 14-Jun | 5 | 125 |
| 21-Jun | 5 | 130 |
| 28-Jun | 5 | 135 |
| 5-Jul | 5 | 140 |
| 12-Jul | 5 | 145 |
| 19-Jul | 5 | 150 |
| 26-Jul | 5 | 155 |
| 2-Aug | 5 | 160 |
| 9-Aug | 5 | 165 |
| 16-Aug | 5 | 170 |
| 23-Aug | 5 | 175 |
| 30-Aug | 5 | 180 |
| 6-Sep | 5 | 185 |
| 13-Sep | 5 | 190 |
| 20-Sep | 5 | 195 |
| 27-Sep | 5 | 200 |

### Supplementary Text S2: Extended model results on effect of NPIs

Figure S1 shows the cumulative attack rate and death rate beginning January 11, 2021 for the NPI scenarios described in Table S5 and 3 different estimates of R_0_ (R_0_=2.4; R_0_=2.8 and R_0_=3.2). In all scenarios involving vaccination, the timeline of vaccination followed the calendar in Table S4. Results in section *Effect of NPIs* in the main text correspond to Scenarios N0 to N5 of Table S5. Without interventions or vaccination (N0) the median national attack rate was 38% for R_0_=2.4, 45% for R_0_=2.8 and 50% for R_0_=3.2. Reductions due to NPIs were qualitatively consistent across the 3 estimates of R_0_, i.e. regardless of R_0_, the scenarios yielding the greatest reduction in infections and deaths nationally were N4, N5, N6 and N9. The best scenarios were therefore those in which NPIs were either first strengthened and then relaxed, or maintained at initial levels until 140 million people were vaccinated. Figure S2 compares the attack rate at the state level for the scenarios considered in the main text (N0 to N5). Among them, Scenario N4 and N5 had the lower attack rate for all states. At the national level, N4 averted more infections and deaths than N5, but in a few states where initial susceptibility was above 74% (Alaska, Vermont, Oregon, New Hampshire, Maine, Hawaii, Washington) N5 was substantially better than N4 (Figure S2).

**Table S5:** All NPIs scenarios considered in the analysis. Scenario N0 and N1 are, respectively, the non-intervention scenario and the only-vaccination scenario. In scenarios N2 to N6, relaxation of NPIs is triggered by milestones (phase completion or 140 million persons vaccinated). In Scenarios N7 to N9, relaxation of NPIs is triggered at specific time points.

| SCENARIO | VACCINATION | DESCRIPTION |
| --- | --- | --- |
| N0 | NO | NPIs immediately relaxed |
| N1 | YES | NPIs immediately relaxed |
| N2 | YES | NPIs maintained until completion of Phase 1a, then immediately relaxed |
| N3 | YES | NPIs maintained until completion of Phase 1a, then gradually relaxed upon completion of (1a,1b,1c) |
| N4 | YES | NPIs strengthened until completion of Phase 1a, then gradually relaxed upon completion of (1a,1b,1c) |
| N5 | YES | NPIs maintained until vaccination of 140 million people, then gradually relaxed in 3 1-month steps |
| N6 | YES | NPIs maintained until vaccination of 140 million people, then immediately relaxed |
| N7 | YES | NPIs as current, then immediately relaxed after 1 month |
| N8 | YES | NPIs as current, then gradually relaxed after 1 month in 5 1-months steps |
| N9 | YES | NPIs strengthened for 1 month then gradually relaxed.in 5 1-months steps |

**Figure S1:**
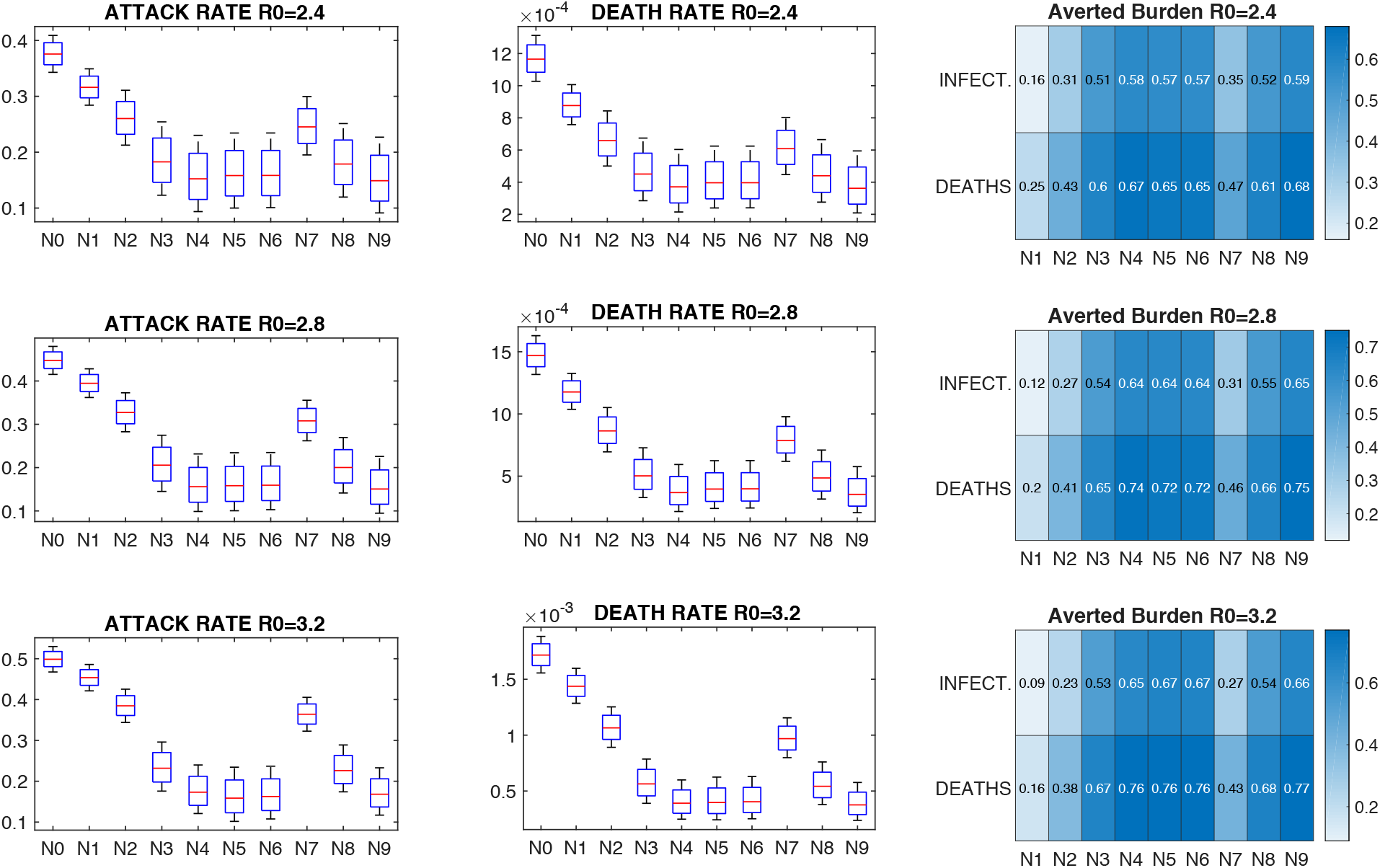
Attack rate and death rate for all NPIs. Cumulative attack rate (first column), cumulative death rates (second column) and averted burden of infection and deaths (third column) for R0=2.4, R0=2.8, R0=3.2. Attack rate and death rate are measured as the fraction of the total population that was infected with SARS-COV-2 and the fraction of the total population that died between January 11 2021 through April 2022.

**Figure S2:**
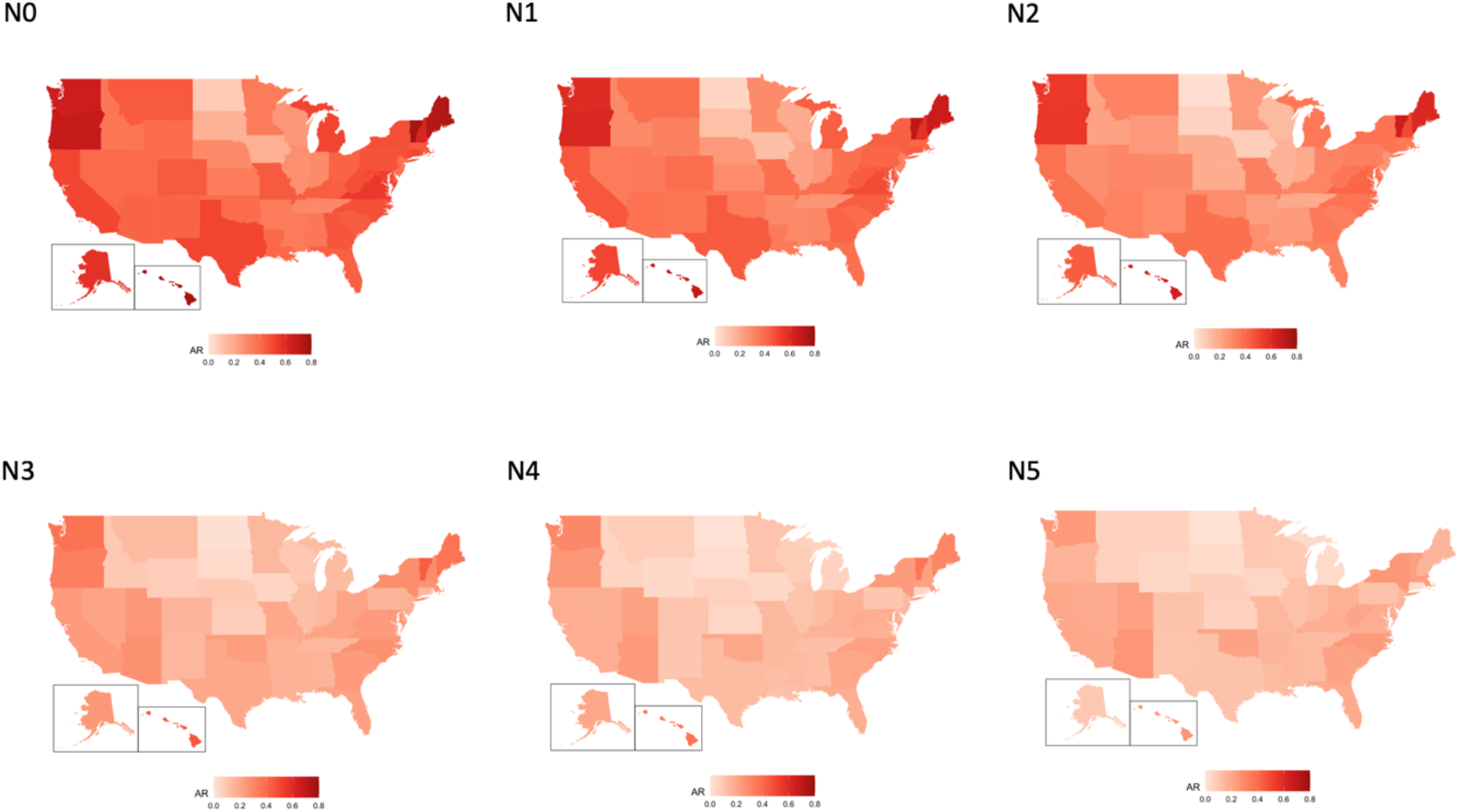
Attack rate by state in scenarios N0 to N5. Maps show the mean estimates of the attack rate (AR) for each state with R_0_=2.8 for the 6 scenarios N0 to N5.

### Supplementary Text S3: Sensitivity Analysis

#### 3.1 Sensitivity to variation of initial susceptibility

The attack rate and death rate largely depended on the estimate of initial population susceptibility. Figure S3 shows the attack rate and death rate when initial susceptibility was increased or decreased 20% from the inferred estimate (65%), corresponding to 78% and 52% initial susceptibility, respectively. The median attack rate in scenario N0 varied from 27% in the decreased susceptibility sensitivity analysis to 62% in the higher susceptibility sensitivity analysis. The ranking of the 9 NPI scenarios of Supplementary Table 5 in terms of averted burden matched the original analysis, with scenarios N4, N5, N6 and N9 yielding the greatest reductions of infections and deaths. However, vaccination in the weaker NPI scenarios had a smaller relative impact when initial susceptibility was higher (Supplementary Figure S3).

**Figure S3:**
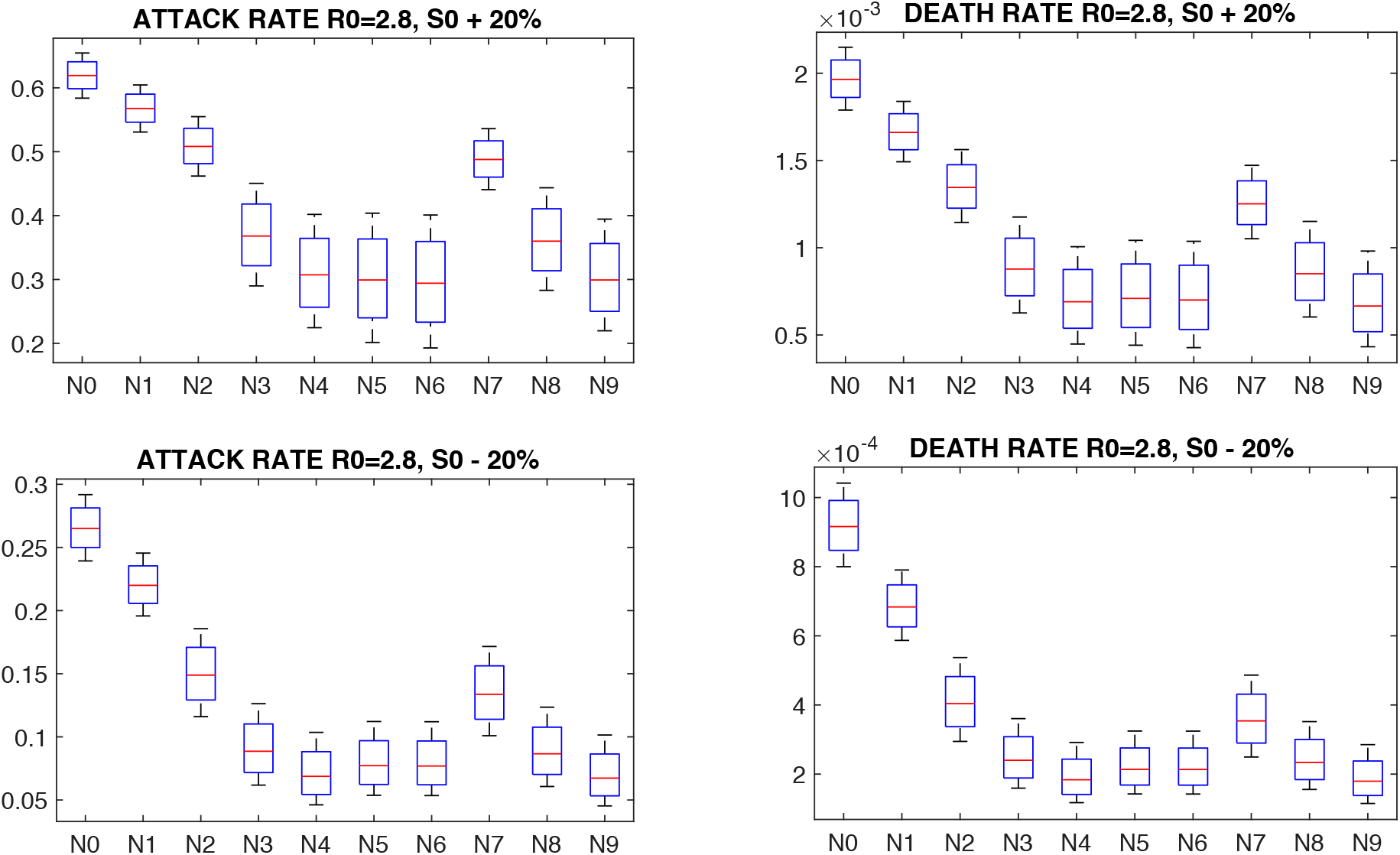
Sensitivity to variation of initial susceptibility. Attack rate and death rate when initial susceptibility in each state is increased by 20% (upper panels) or decreased by 20% (lower panels). Here, R_0_=2.8, the vaccination calendar is shown in Table S4, and coverage is baseline c (80% HC, 70% for population ≥65 and adults with RF and 60% for others up to availability).

#### 3.2 Sensitivity to different estimates of vaccine efficacy

The estimate of vaccine efficacy used in the model was based on results from the Pfizer and Moderna phase 3 trials (*4, 5*). However, experimental error (e.g. PCR sensitivity) and possible reduction of effectiveness due to new emerging viral variants (*20*) could impact the actual efficacy of the vaccine. When efficacy was reduced by 20% (both after the first and second dose), the averted infections and deaths were 70-95% and 75-98% of original analysis, respectively (Supplementary Figure S4).

**Figure S4:**
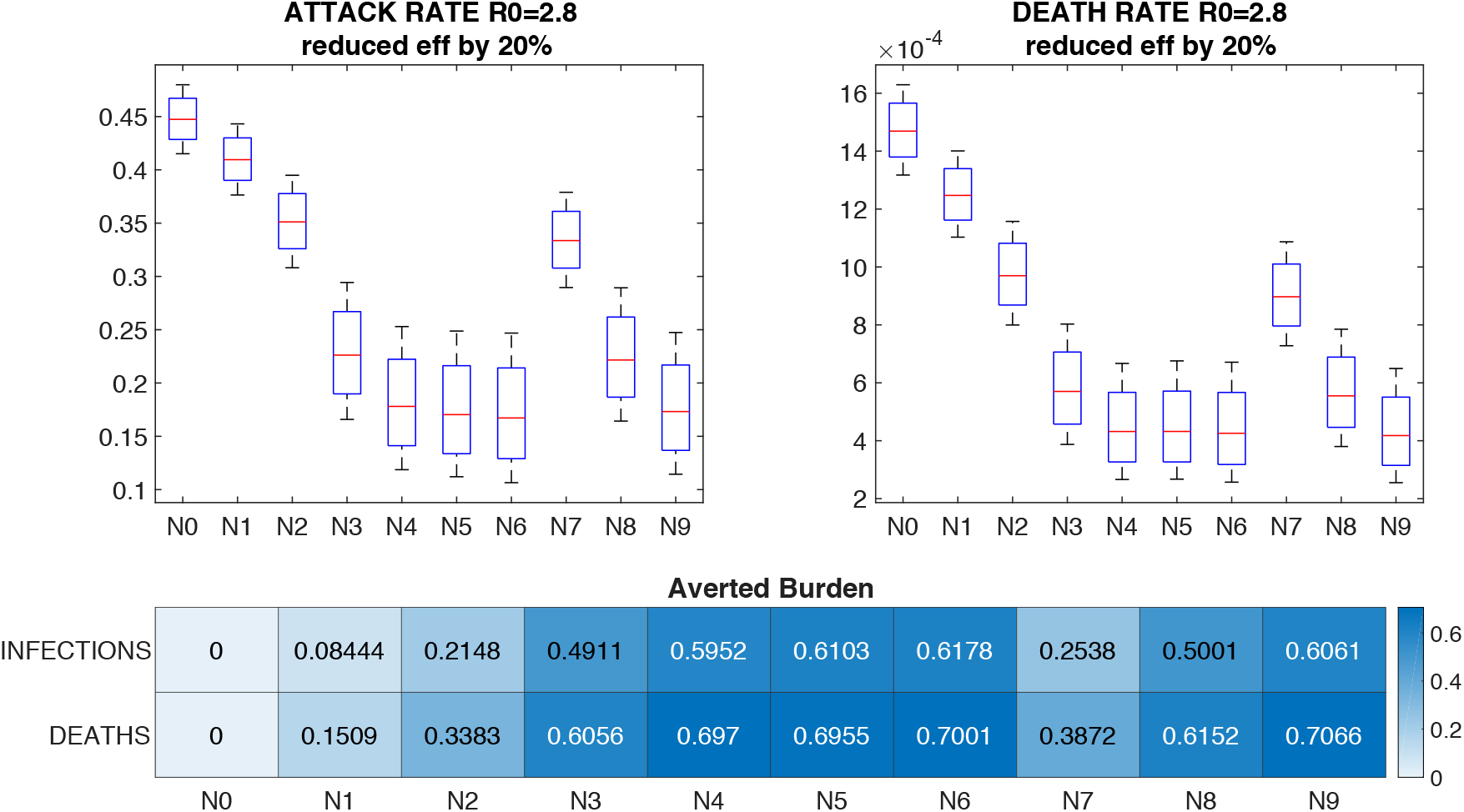
Attack rate and death rate for vaccine efficacy reduced by 20%: (*ϵ*_1_ = 0, *ϵ*_2_ = 72% *ϵ*_3_ = 76%). Here R_0_=2.8, the vaccination calendar is presented in Table S4, and coverage is baseline c (80% HC, 70% for population ≥ 65 and adults with RF and 60% for others up to availability).

#### 3.3 Sensitivity to variation of reduction in contact rates

We tested the effect of varying the relative reduction of contact rates in matrix M of Supplementary Table S2 among population groups. The overall transmission rate was determined from the estimate of R_t_ for each state, derived from the metapopulation model, and by the NPIs scenario adopted; however, relative ratios of NPI-induced contact reduction to the POLYMOD contact rates (*30*) in different groups could be tuned. The values used in simulations were a 60% contact reduction in school, 60% contact reduction at work, no contact reduction at home and 30% contact reduction in other settings (Supplementary Table S2). Altering these percentages yielded less than 2% difference in attack rate but had a more substantial effect on deaths. However, the qualitative results were unchanged (Supplementary Figure S5).

**Figure S5:**
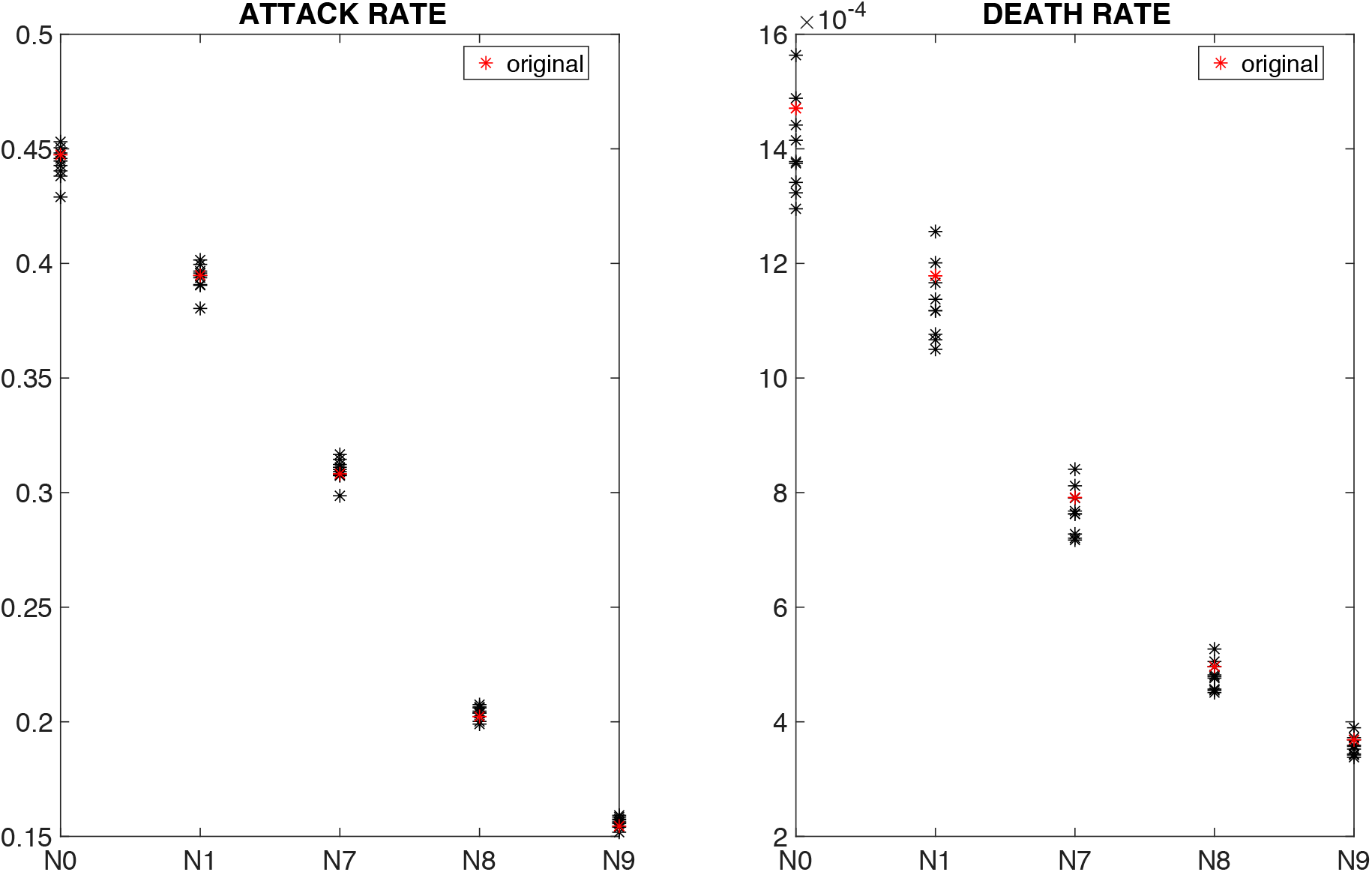
Sensitivity of the attack rate to varying the relative ratios of contact rates. Results are for R_0_=2.8, the vaccine calendar in Table S4 and coverage baseline c (80% HC, 70% for population ≥65 and adults with RF and 60% for others up to availability). Symbols correspond to mean estimates of the attack rate and death rate corresponding to different choices of relative contact reduction due to NPIs. Red symbols are the mean estimate for the original setting (60% contact reduction in school, 60% contact reduction at work, no contact reduction at home, and 30% contact reduction in other settings). Black symbols are the mean estimates for 10 arbitrary perturbations of the relative contact reduction at school, work and other settings.

#### 3.4 Single dose vaccination

We compared the performance of 2-dose vaccination with a 1-dose vaccination campaign based on the same cumulative vaccine availability (400 million doses total). We set the efficacy of 1-dose vaccination to 90% 12 days after administration (Table S3). The population vaccinated per week increased from 5 to 10 million, and the stock was sufficient to vaccinate the entire population as second doses were not administered and all available vaccines were used as first doses. We analyzed two different uptake scenarios: the baseline scenario *c* (80% HC, 70% risk groups, 60% others, for a cumulative maximum uptake of 64%) and scenario *c*_*99*_ where coverage was 99% in all groups. For both coverage scenarios, the one-dose vaccination yielded more averted infections and deaths than the two-dose vaccination for all the NPI settings considered (N1, N7, N8, N9). Specifically, with uptake scenario *c*, there were 6-12% more averted infections and 5-15% more averted deaths depending on the NPI scenario (Figure S6). With uptake scenario *c*_*99*,_ in which not only the allocation speed, but also the total population covered was significantly increased in the 1-dose vaccination campaign, there were 7-13% more averted infections and 5-17% more averted deaths (Figure S7) with respect to scenario N0.

**Figure S6:**
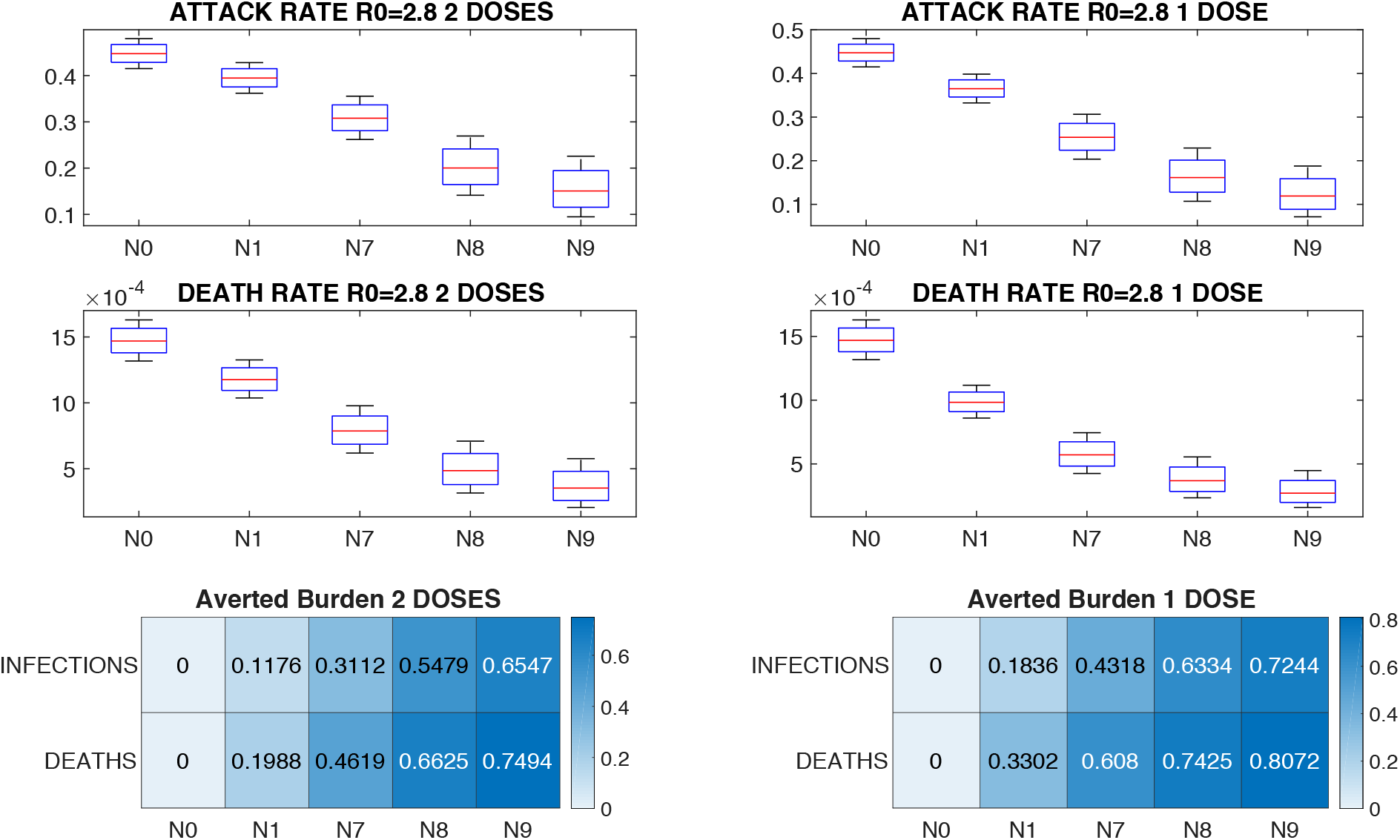
Comparison between one-dose and two-doses vaccination in uptake scenario c. Left panels show the attack rate, death rate and averted burden for two-dose vaccination with 61.5% coverage and 5 million vaccination per week; right panel show the attack rate, death rate and averted burden for one-dose vaccination (right) with 64% coverage and 10 million vaccination per week, in uptake scenario *c*. The NPI scenarios N0, N1, N7, N8, N9 are described in Table S5.

**Figure S7:**
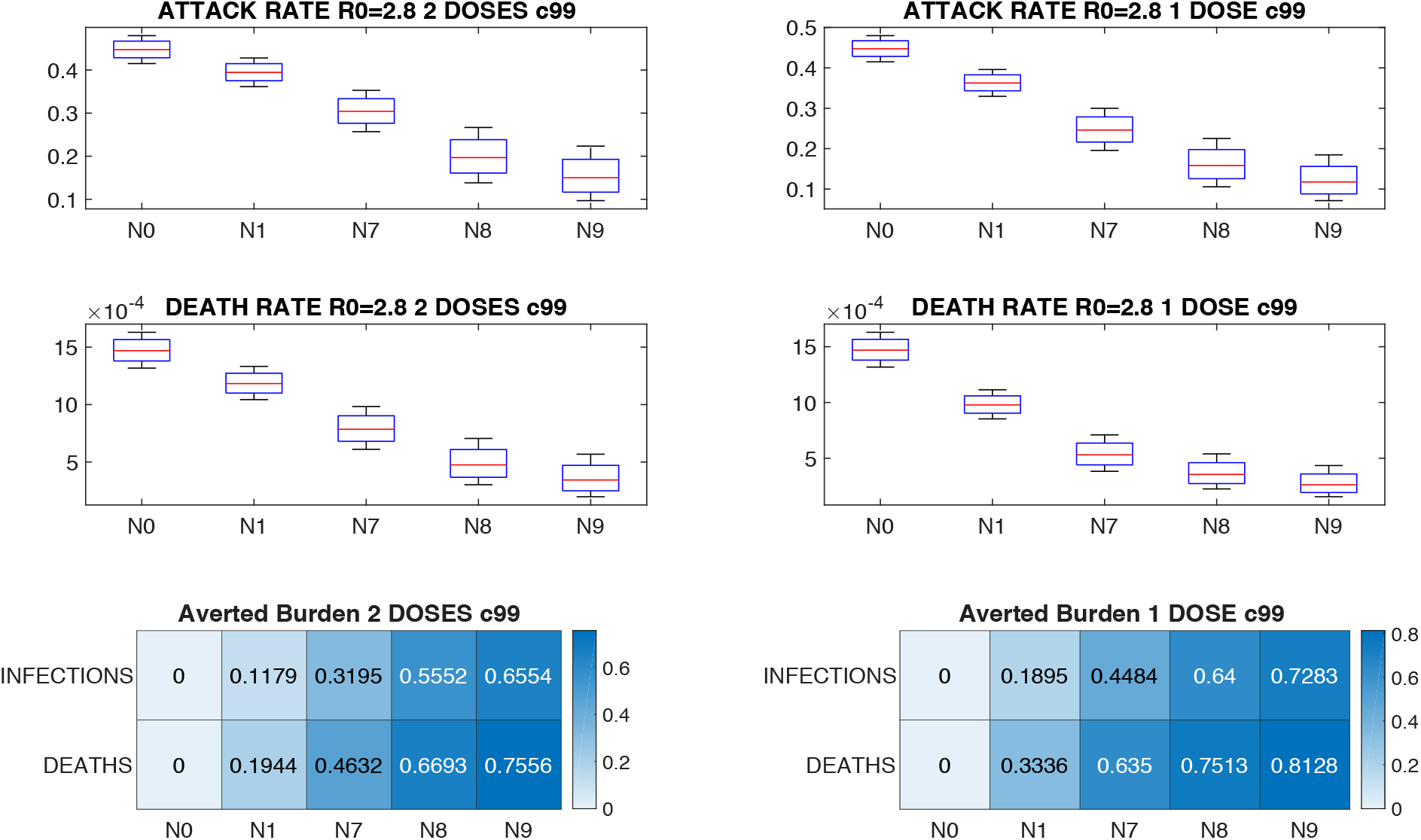
Comparison between one-dose and two-doses vaccination in uptake scenario c_99_. Left panels show the attack rate, death rate and averted burden for two-dose vaccination with 61.5% coverage and 5 million vaccination per week in uptake scenario c_99_; right panel show the attack rate, death rate and averted burden for and one-dose vaccination (right) with 99% coverage and 10 million vaccination per week. The NPI scenarios N0, N1, N7, N8, N9 are described in Table S5.

#### 3.5 Disease-blocking vaccine versus infection-blocking vaccine

Vaccine trials demonstrated a 95% reduction of SARS-COV-2 disease in vaccine recipients. However, it is currently unclear whether the vaccine prevents infection or symptomatic disease upon infection. Throughout the analysis we assumed the former, by multiplying the transmission rate by the reduction constant *ε*. Here, we repeat the analysis assuming that vaccination does not prevent infection (*ε* = 1), and assumes instead that the Exposed Vaccinated in system 1) are 95% less likely to develop a symptomatic infection 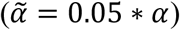 than the Exposed (unvaccinated). The first panel of Figure S8 shows the averted burden of the disease-blocking vaccine for R_0_=2.8 for all NPI scenarios in Table S5. Overall, averted infections were substantially reduced: averted infections in the disease-blocking vaccine were 1/3 of infection-blocking vaccine in some scenarios, whereas averted deaths were 85% to 95% of the infection-blocking vaccine scenarios.

**Figure S8:**
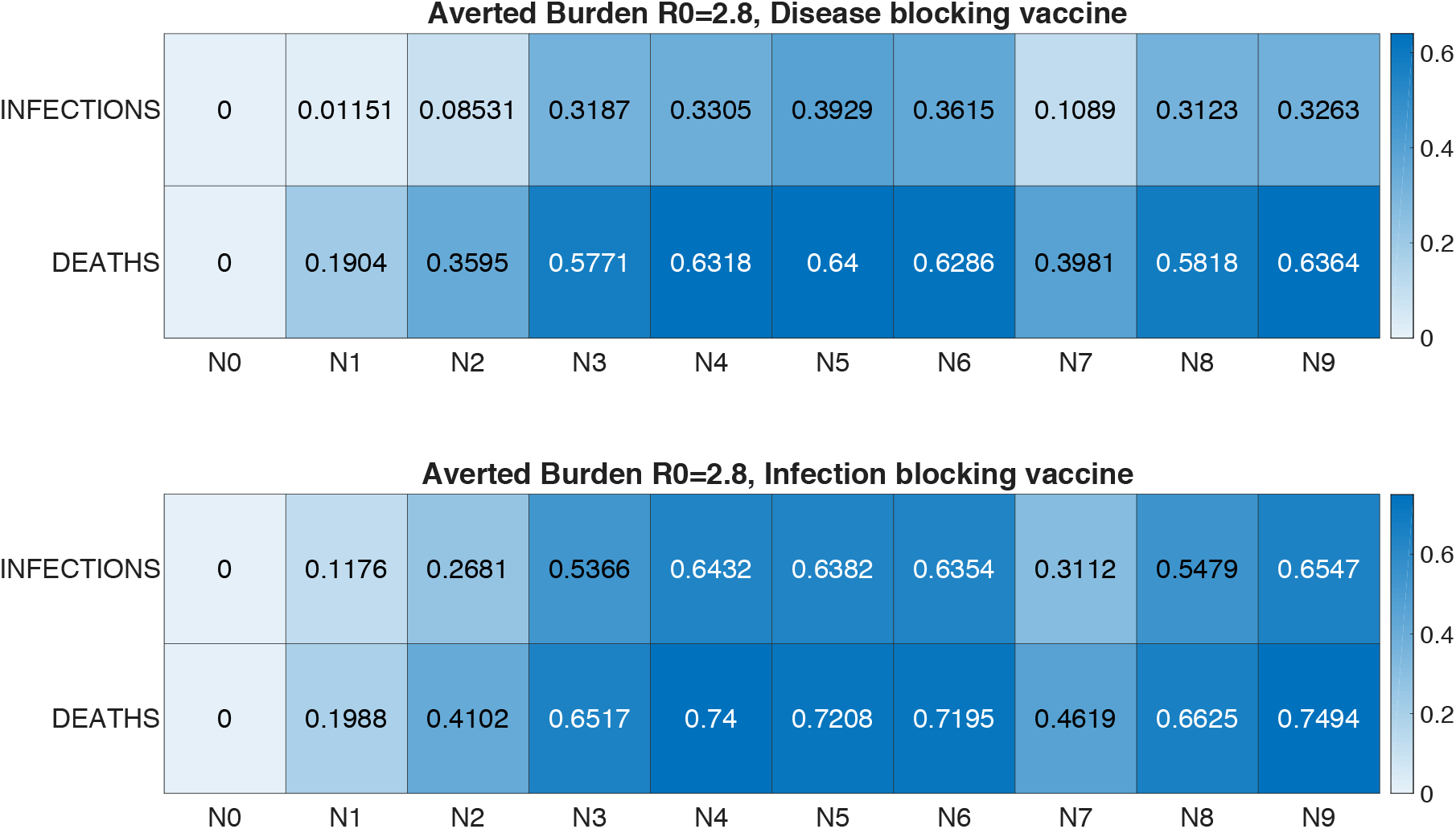
Comparison between attack rate and death rate for disease-blocking vaccination and infection-blocking vaccination. Here, R_0_=2.8, vaccination calendar is as presented in Table S4 and coverage is baseline c (80% HC, 70% for population ≥65 and adults with RF and 60% for others up to availability).

### Supplementary Text S4: Extended model results for the effect of allocation rate

Figure S9 extends the results of Figure 4 in the main text depicting the effect of allocation rate for 3 levels of R_0_. The qualitative behavior was similar across the different choices of R_0_: increasing the weekly deployment from 5 to 11 million doses yielded 7-14% more averted infections and 7-17% more averted deaths for R_0_=2.4; 9-16% more averted infections and 7-18% more averted deaths for R_0_=2.8; and 7-16% more averted infections and 8-20% more averted deaths for R_0_=3.2 with respect to scenario N0.

**Figure S9:**
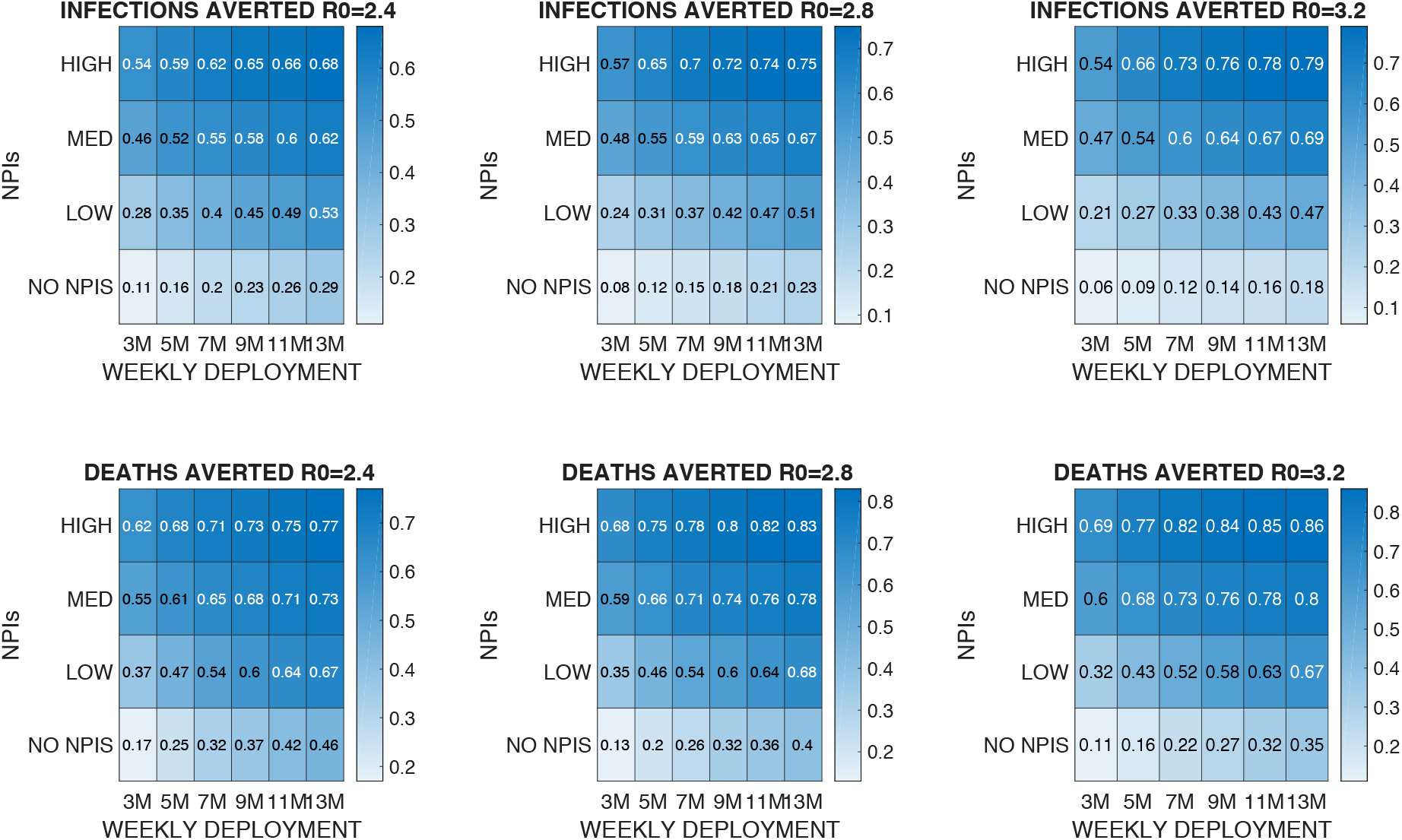
Averted burden for varying administration rates. Panels show the fractional averted infections and deaths for the 4 time-triggered NPI scenarios N1(NO NPIs), N7 (LOW), N8 (MED), and N9 (HIGH) for 3, 5, 7, 9, 11 and 13 million individuals vaccinated each week, relative to the baseline scenario (N0). Each column displays the results corresponding to a different R_0_.

**Figure S10:**
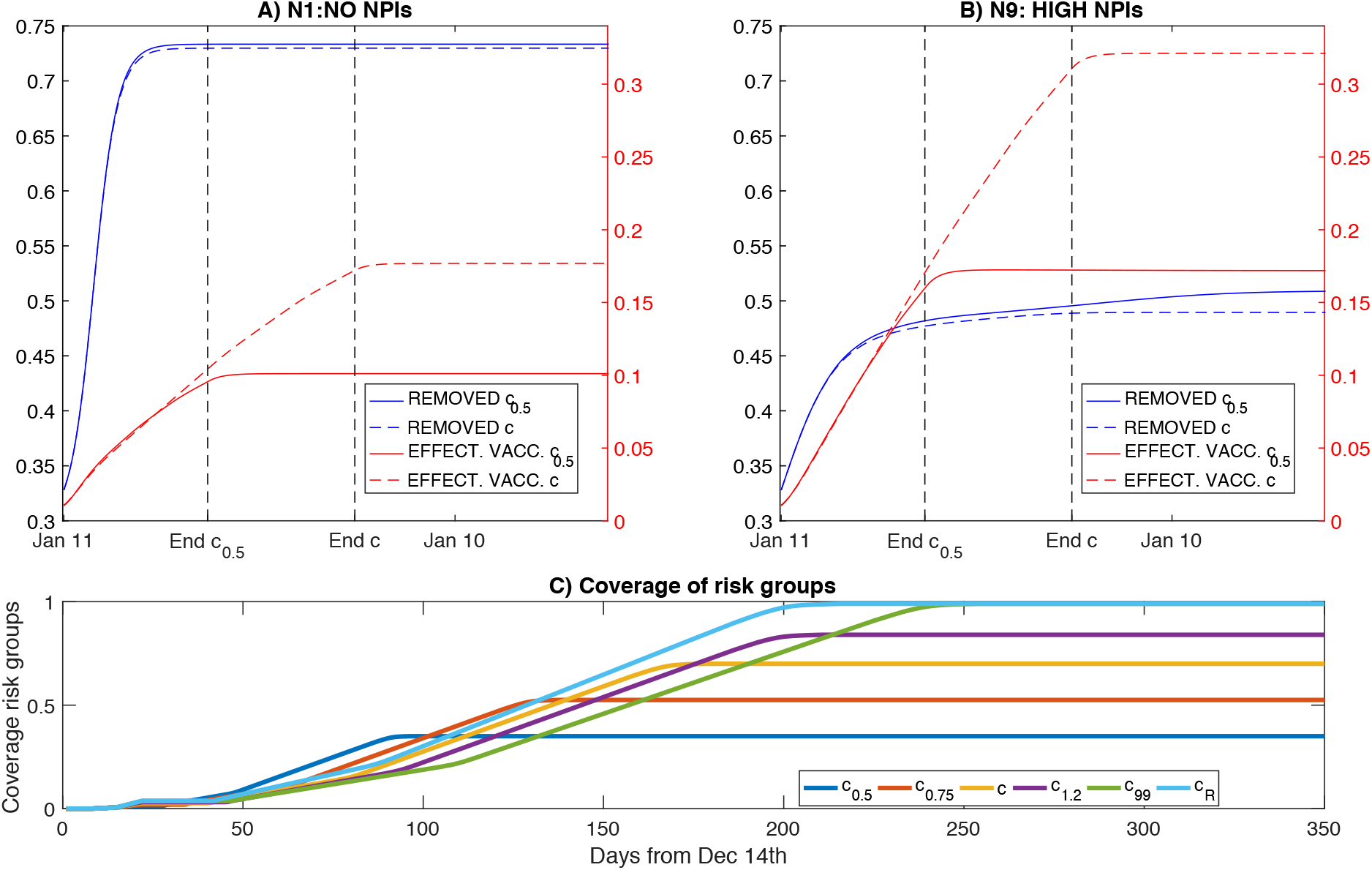
Effect of NPIs and vaccination uptake on population immunity. Panels A and B) Cumulative individuals no longer susceptible or infected (blue lines: recovered + deceased) and effectively vaccinated (red lines: recipients of vaccine who were susceptible) in scenario N1 (NO NPIs, panel A) and N9 (HIGH NPIs, panel B). Dashed blue and red lines refer to uptake scenario c and solid blue and red lines refer to uptake scenario c_0.5._ Black vertical dashed lines mark the end of vaccination (first doses) in c and c_0.5._ Panel C) Vaccine coverage of risk groups (adults with RF and population ≥65 with and without RF) through time from December 14, 2020 for the 6 uptake scenarios (c_0.5_, c_0.75_, c, c_1.2_, c_99_, c_R_).

### Supplementary Text S5 Extended model results on the effect of vaccine uptake

Figure S11 extends the analysis of Figure 5 in the main text testing the effect of population uptake of vaccine for 3 levels of R_0_. The limited effect of uptake is consistent regardless of R_0._ Coverage scenario c_R_ yields the greatest reduction in deaths for all NPI scenarios and for all choices of R_0_. This moderate effect was not dependent on the cumulative number of doses: when we increased total available doses from 400 million to 600 million (from 200 million vaccinated to 300 million), and maintained an administration rate of 5 million vaccinations per week, the attack rate decreased by at most 0.003% in the stronger NPI scenarios and did not vary in the weaker NPI scenarios.

**Figure S11:**
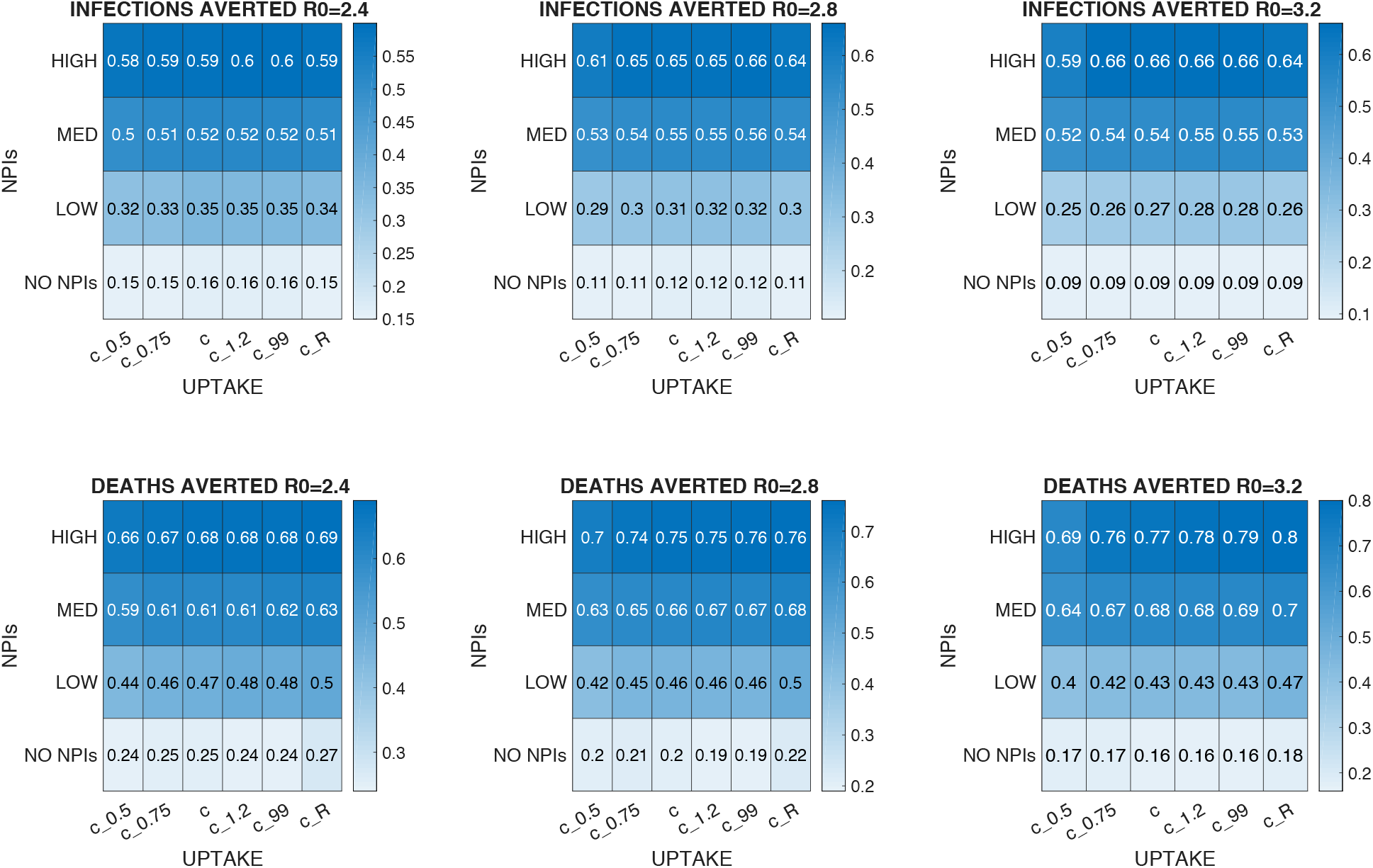
Averted burden for varying uptake scenarios. Panels show the fractional averted infections and deaths in the 4 time-triggered NPI scenarios N1 (NO NPIs), N7 (LOW), N8 (MED), N9(HIGH) for the 6 choices of population coverage (c_0.5_, c_0.75_, c, c_1.2_, c_99_, c_R_), 400 million total available doses, and 3 levels of R_0._ All numbers represent the fractional reduction relative to the baseline scenario (N0).

